# Empirical limitations of current low-intensity focused ultrasound simulation platforms

**DOI:** 10.64898/2026.01.20.26344460

**Authors:** Sierra D. Brandts, Brooke R. Staveland, Ananya Madhusudhan, Ryan T. Ash, Tommaso Di Ianni, Leo P. Sugrue, Andrew D. Krystal, Khaled Moussawi

## Abstract

**Background:** Transcranial low-intensity focused ultrasound (LIFU) is an emerging tool for noninvasive neuromodulation, with potential therapeutic applications across a range of neuropsychiatric disorders. As the field advances toward clinical translation, individualized simulations of sonication parameters are critical for estimating dosing to ensure safety, efficacy, and reproducibility. Currently, k-Plan (Brainbox Ltd.) and BabelBrain (open-source) are two widely used software packages for sonication simulation, yet their outputs have not been empirically compared. While CT scans remain the gold standard for capturing fine anatomical details of the skull, their use carries radiation risk. As an alternative, algorithms generating MRI-based pseudo-CTs have been developed to approximate skull bone properties.

**Objective:** In this study, we directly evaluated the consistency of simulation results from the two platforms, k-Plan and BabelBrain. We also examined the impact of skull modeling using CT vs. pseudo-CT on LIFU simulation results.

**Methods:** We compared LIFU simulation results between the two platforms when applied to the same individuals, trajectories, and using the same sonication protocol parameters. To assess the validity of using pseudo-CTs for simulations, we also compared simulation outputs generated using CT versus pseudo-CT inputs.

**Results:** Our results reveal substantial discrepancies both between software platforms (k-Plan vs. BabelBrain) and between skull modeling approaches (CT vs. pseudo-CT). These findings underscore the urgent need for ground-truth validation of current sonication simulation software and highlight the importance of further optimization of pseudo-CT algorithms before widespread clinical adoption.

## Introduction

Transcranial low-intensity focused ultrasound (LIFU) is an emerging technology that enables noninvasive modulation of both superficial and deep brain structures, thereby overcoming a major limitation of other noninvasive neuromodulation tools [1-3]. LIFU has been applied to a variety of brain targets, including the anterior cingulate cortex, subgenual cingulate cortex, insula, amygdala, ventral striatum, and other deep brain structures, in both healthy volunteers and patients with neuropsychiatric disorders [4-11]. With the number of clinical trials steadily increasing, ensuring accurate and reproducible dosing has become a central challenge, especially in light of the recent report of a serious adverse event during the delivery of a presumed low-intensity focused ultrasound sonication protocol, underscoring the importance of accurately simulating in situ pressures and temperatures [12].

Accurate dosing is essential for safety, efficacy, and reproducibility. At sufficiently high intensities, ultrasound pressure waves may cause tissue damage through mechanisms such as thermal necrosis or cavitation. While the U.S. Food and Drug Administration (FDA) has not yet issued guidelines specific to LIFU, most research groups follow existing diagnostic ultrasound safety guidelines and expert consensus recommendations (e.g., the International Transcranial Ultrasonic Stimulation Safety and Standards consortium (ITRUSST)) [13]. Key safety measures include the spatial-peak pulse-average intensity (I_SPPA_), the mechanical index (MI), and the maximum allowable temperature rise. For a given sonication protocol, in-situ parameters are strongly influenced by the skull’s biophysical properties, which vary considerably across individuals and across cranial regions within the same individual [14]. Such variability affects ultrasound propagation, focusing accuracy, and ultimately the delivered dose at the target.

To address these challenges, personalized simulations have become a cornerstone of LIFU research, a key step forward in safety and accuracy for the field of ultrasonic neuromodulation. These simulations estimate ultrasound propagation and focal pressure fields based on individual anatomical data, most commonly from CT imaging, which provides high-resolution characterization of skull thickness and density. However, because of radiation exposure, there is increasing interest in generating pseudo-CTs from MRI sequences to approximate skull properties without ionizing radiation [15]. Whether such approaches provide sufficient accuracy for clinical or research applications remains an open question.

Several computational tools have been developed to perform such personalized simulations. Two widely adopted platforms are k-Plan (BrainBox Ltd.) [16], a commercial software built on the *k-Wave* acoustic simulation toolbox and executed via cloud-based computing [17], and BabelBrain, an open-source platform recently integrated with the Rogue Research Brainsight neuronavigation system, which operates via local computation [18]. Despite their growing use, these two platforms have not been directly compared.

In this study, we address these gaps by systematically evaluating the consistency of simulation outputs between k-Plan and BabelBrain when applied to the same individuals, sonication trajectories, and protocol parameters. We also discuss the different modeling assumptions and sources of discrepancies between the simulation platforms. We further assess the impact of input modality by comparing simulations derived from CT versus pseudo-CT skull models. By identifying discrepancies and potential sources of variability, our findings aim to inform best practices for LIFU sonication planning, highlight the need for standardized validation methods, and guide optimization of pseudo-CT algorithms for safer and more reproducible neuromodulation.

## Methods

### Sample Demographics

Imaging data from two groups of subjects (group 1, N = 8; group 2, N = 5) were used for the two experiments. De-identified CT and MRI imaging data were originally acquired for clinical care purposes or as part of other ongoing studies. All subjects had provided written informed consent to allow using their de-identified scan data as part of the relevant research protocol approved by the Institutional Review Board at the University of California, San Francisco.

### Image Acquisition

Structural MRI (GE SIGNA Premier scanner), including T1- and T2-weighted sequences (T1w and T2w, respectively), and CT imaging were acquired from all eight subjects. T1w images were acquired using an MPRAGE sequence; variations in acquisition parameters occurred across subjects due to scanner protocol differences across the subject groups (TR = 1520–2800 ms, TE ≈ 2.5 ms, TI = 800–1000 ms, flip angle ≈ 9°) and T2w images were acquired using spin echo (SE) sequence (TR = 2700–4000 ms, TE = 90–160 ms, and echo train lengths of 130–250, flip angle = 90°). The CT slice thickness was 0.625 to 1.0 mm and pixel spacing ranged from 0.48 to 0.59 mm. All scans were reconstructed using bone windows and the BONEPLUS convolution kernel, ensuring high bone anatomical detail. Zero-TE (ZTE) sequences were obtained from five subjects. The ZTE sequences were used only to compare simulation outputs between ZTE and CT images, and were not used to compare the different simulation platforms. ZTE parameters consisted of TR/TE = 530/0.01 s, 1° flip angle, a pixel bandwidth of 244 Hz/pixel, and single echo, which corresponds with the settings used in Miscouridou et al. [15]. The PURE/SCENIC setting was applied for chemical shift reduction and signal homogenization.

### Image preprocessing

All raw MRI and CT images were first converted from DICOM to NIfTI format. This preprocessing pipeline is illustrated in Figure 1. An isotropic voxelization was then performed at 1 mm resolution on the T1-weighted (T1w) and T2-weighted (T2w) images using FSL FLIRT (version 6.0.7.13). To ensure consistent spatial orientation between k-Plan and the Brainsight neuronavigation system [19], the T1w and T2w images were resliced. Reslicing is required to prevent the transducer and target positions from being misaligned across platforms; without reslicing, Brainsight reformats and reorients images to be squared within each viewing plane, whereas k-Plan displays the images in their acquired orientation. T1w and T2w images were segmented and reconstructed using the SimNIBS head modeling *charm* tool [20], which generates anatomical head models used as the inputs for BabelBrain simulations [21]. CT and ZTE NIfTI files were not preprocessed prior to simulations to preserve the skull mask and avoid smoothing effects introduced by isotropic resampling or normalization during co-registration. Changes to the skull mask and smoothing alter Hounsfield units (HU) across voxels, thereby affecting simulation accuracy. The CT images were retained at their native acquired resolution (0.48–0.59 × 0.48–0.59 × 0.625–1.0 mm³).

**Figure 1.**
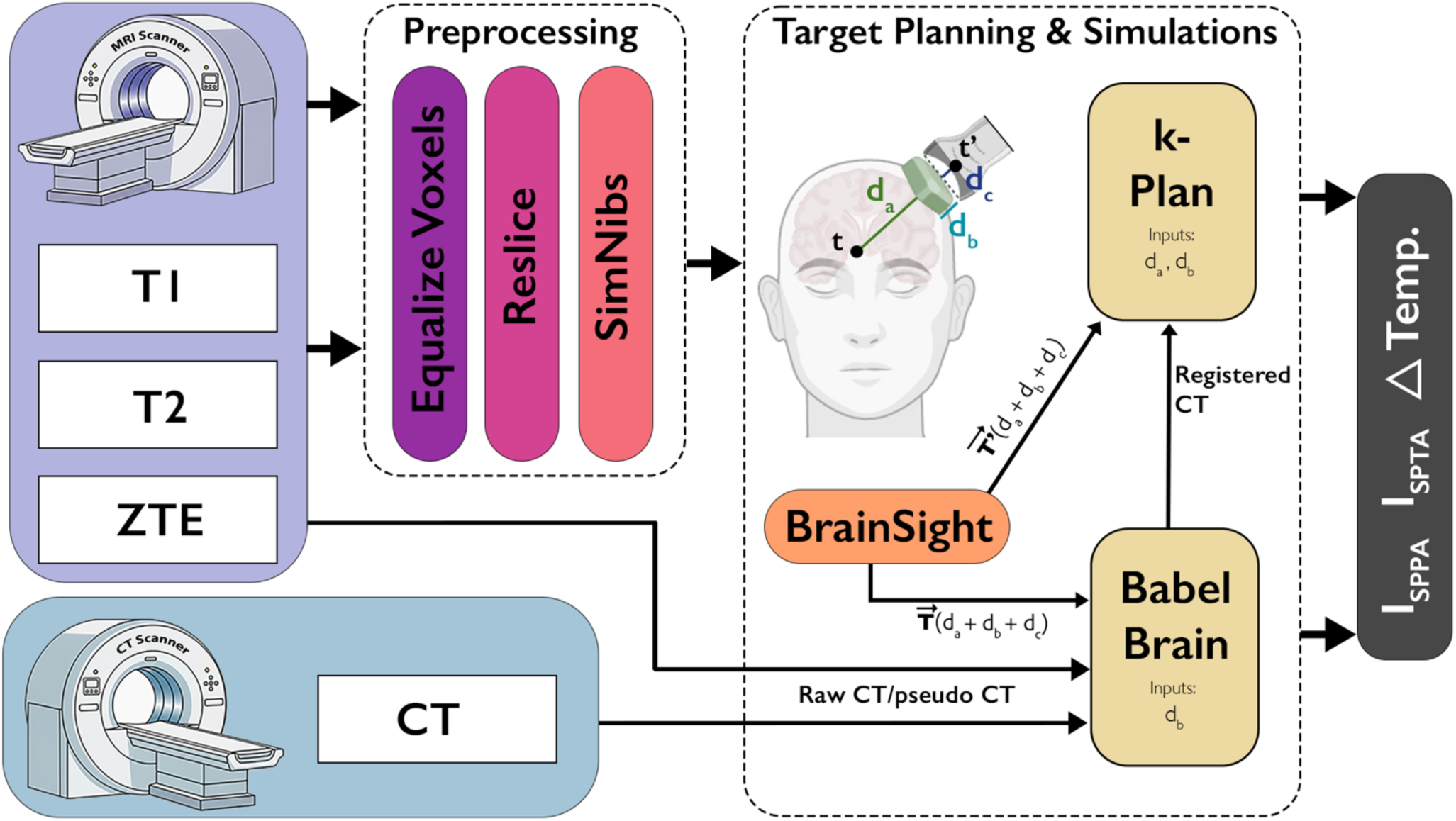
Overview of imaging, preprocessing, target-planning, and simulation workflow. Structural MRI (T1w, T2w, ZTE) and CT scans were acquired and converted to NIfTI format. Only T1w and T2w images were preprocessed (voxel standardization, reslicing, SimNIBS segmentation) to ensure consistent visualization across Brainsight and k-Plan. CT and ZTE volumes were kept at native resolution to avoid smoothing prior to acoustic simulations. Registered CTs generated in BabelBrain were imported directly into k-Plan. Target planning for both platforms was performed in Brainsight. Although the beam trajectories were identical across the two platforms, k-Plan expects the trajectory to be anchored to the transducer position (t’), while BabelBrain expects the trajectory to be anchored to the brain target (t). To account for this difference, two trajectory files were created in Brainsight. First, 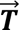 was created for BabelBrain, which anchors the trajectory to t. Within BabelBrain, the distance between t and the scalp (d_a_) was automatically computed using the CT and the distance between the transducer exit plane and the transducer apex (d_c_) was accounted for by specifying the transducer model. The coupling gel creates an additional distance (d_b_) between the scalp and the transducer exit plane, and this distance was manually added in BabelBrain using the ‘TPO distance’ and ‘Distance Tx outplane to skin’ fields. To create an equivalent trajectory 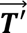 for k-Plan in Brainsight, we add the three offsets (d_a_, d_b_, d_c_) to the Crosshair Origin field in Brainsight to find t’ for the k-Plan simulation. In k-Plan, d_a_, as calculated by BabelBrain, and d_b_ were added to the ‘Focal Distance’ field. d_c_ is similarly accounted for in k-Plan by selecting the transducer model. Simulation outputs analyzed included max and target I_SPPA_, I_SPTA_, and maximum change in skull temperature.

### Target Planning

Target planning for both BabelBrain and k-Plan simulations was performed using Brainsight (version 2.5.5). A SimNIBS project was generated for each subject, and the ventral striatum (VS) was selected as the primary target region of interest. The VS was identified for each subject as the part of the striatum where the caudate and putamen converge ventrally, anterior to the anterior commissure and inferior to the internal capsule. In a supplementary analysis, the ventral anterior insula (vAI) was selected as a different target region of interest. The vAI was identified for each subject as the insular cortex anterior to the central insular sulcus. The target was further defined as the most anterior and ventral extent of the insula with a slight medial adjustment within the gray matter to ensure appropriate depth for the CTX-250-4CH transducer.

To examine how skull geometry and incidence angle influence simulation results, four distinct trajectories from the scalp to the VS were defined and simulated for each subject. Trajectories A and B were selected to be reproducible across subjects regardless of individual skull anatomy by anchoring the transducer at fixed orientations relative to the midsagittal plane (90° for Trajectory A and ∼60° for Trajectory B; Figure 2A–B). These standardized placements ensured consistent skull-entry locations across subjects, but they often intersected subject-specific regions of bone irregularity, frequently producing diffracted focal beams. Trajectories C and D were individualized for each subject by using their CT-derived skull masks to identify the most orthogonal paths relative to the skull surface. Because energy loss from reflection and absorption increases with greater incidence angles relative to the skull [22], BabelBrain and k-Plan may produce more consistent predictions when the transducer is more orthogonal to the skull. Thus, Trajectory C was chosen to represent the most orthogonal ipsilateral trajectory to target. However, in all subjects, a more optimal contralateral Trajectory D could be identified as more orthogonal than Trajectory C (Supplementary Figure 1), and as such this was included in our comparisons. Trajectories were optimized and verified using Brainsight’s Inline and Inline-90 visualization views.

**Figure 2.**
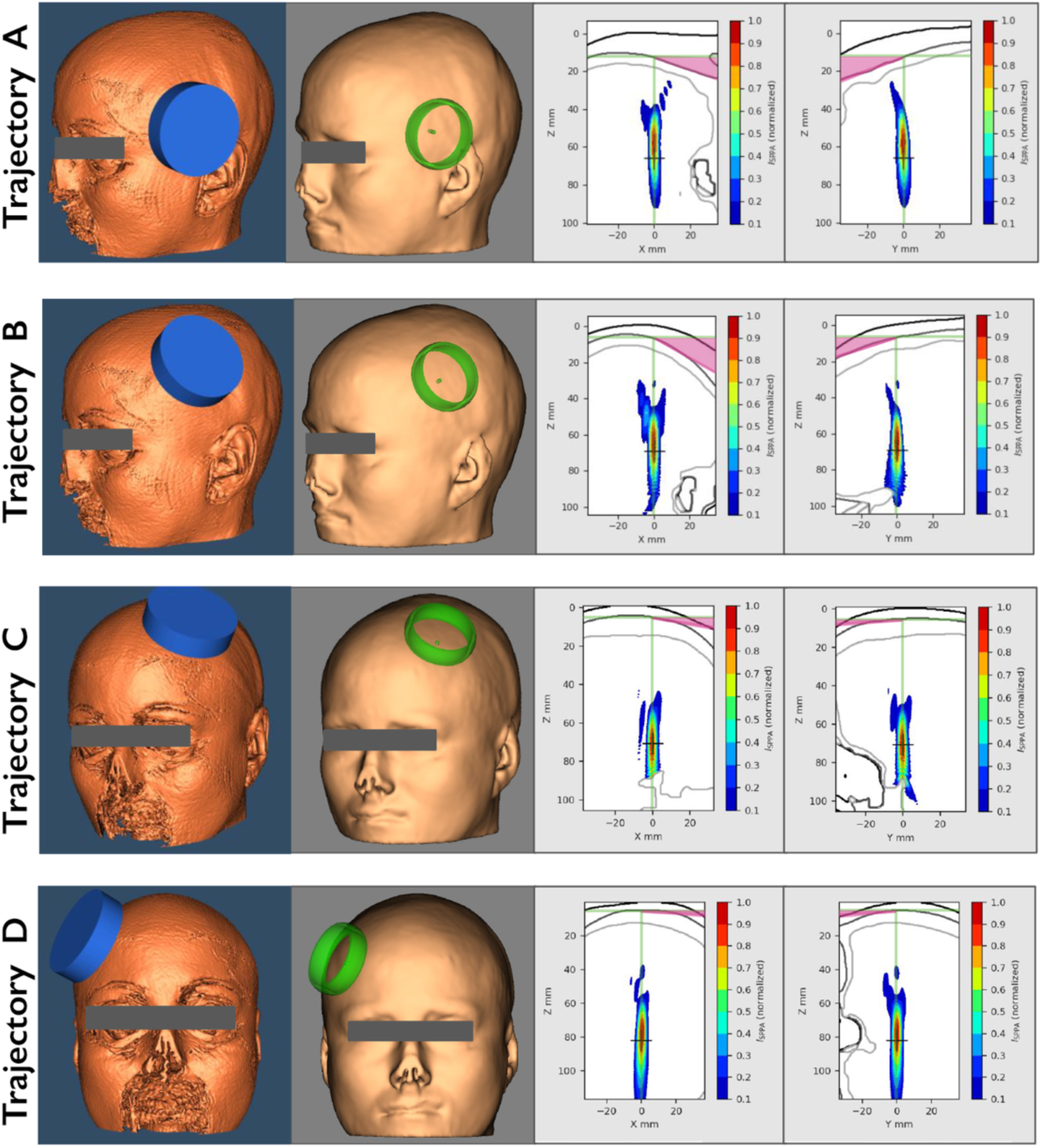
Simulated sonication trajectories. The following four trajectories targeting the ventral striatum were used in all five subjects. Illustrated example trajectories targeting the left ventral striatum from Subject 1. 3D head renderings illustrate the placement of the transducer in k-Plan (panels with blue rings) and Brainsight (panels with green rings), with the green/blue ring indicating the transducer location. For each trajectory, the accompanying 2D maps from BabelBrain display the normalized acoustic intensity along the X–Z and Y–Z planes. In each 2D map, the top black line represents the scalp, the middle black line represents the outer surface of the skull, and the bottom line represents the inner surface of the skull. The pink shading marks the deviation of the ultrasound beam from orthogonality, approximated through a tangent to the skull surface at the center of the ultrasound beam. Trajectory A was perpendicular to the midsagittal plane. Trajectory B was ∼60 degrees from the midsagittal plane. Trajectory C was the most orthogonal trajectory ipsilaterally. Trajectory D was the most orthogonal trajectory, contralaterally – note that trajectory D provided the most orthogonal incidence angle relative to the skull for targeting the ventral striatum across all participants.

BabelBrain expects trajectories to be anchored to the brain target, whereas k-Plan expects a trajectory to be defined with respect to the apex of transducer. To ensure consistent target planning across both platforms, we defined a common trajectory based on the spatial positions of both the target and the transducer. The distance between the target and the transducer along this trajectory is parameterized using three offsets. The first offset, d_a_, represents the target-to-scalp distance. The second offset, d_b_, represents the distance between the scalp and the transducer exit plane and accounts for any coupling medium, such as a gel pad or water bladder, as well as hair or any other substance between the exit plane and the skin. The third offset, d_c_, represents the distance from the transducer exit plane to the apex of the transducer’s curvature (Figure 1).

For BabelBrain simulations, we defined a trajectory 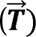 in Brainsight by selecting the target location (t) and then adjusting the anterior–posterior and lateral sliders to establish the desired trajectory between the target and transducer. This trajectory was then exported to the BabelBrain interface, which then calculates the distance between target (t) and the apex of the transducer (t’) using the three offsets. The d_a_ parameter is automatically computed by BabelBrain and is displayed in the ‘Transducer Power Output (TPO) Distance’ field. d_b_ is manually set using the ‘TPO distance’ and ‘Distance Tx outplane to skin’ fields and represents the distance between the scalp and the transducer exit plane. This distance should include any coupling medium, such as a gel pad or water bladder, as well as hair or any other substance between the exit plane and the skin. d_c_ is predefined based on the selected transducer model and does not require user input (d_c_ can be found in transducer manual).

For k-Plan simulations, we use the same trajectory. However, k-Plan expects a trajectory anchored to the transducer apex, which we call 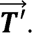 To create 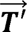 in Brainsight, we take 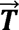 and manually add d_a_, d_b_, and d_c_using Brainsight’s *crosshair offset* slider. d_a_ is pulled from BabelBrain’s ‘Transducer Power Output (TPO) Distance’ field, d_b_ is set manually. This trajectory, which is now anchored to the apex of the transducer, is then exported to the k-Plan interface. Within k-Plan, we then set the ‘Focal Distance’ field to the sum of d_a_ and d_b_. As with BabelBrain, d_c_ is automatically accounted for when the appropriate transducer file is selected. This approach, outlined in Figure 1, converts BabelBrain’s target-based trajectory into a geometrically equivalent transducer-based trajectory for use in k-Plan.

### BabelBrain Simulations

Simulations were performed using BabelBrain version 4.1. The DPX-500-4CH four-channel annular array transducer (active diameter: 64 mm; maximum focal depth: 150 mm) was used for all VS simulations at the default grid resolution of six points per wavelength. The CTX-250-4CH annular array transducer (active diameter: 64.5 mm; maximum focal depth: 63.2 mm) was used for all Anterior Insula simulations at the default grid resolution of six points per wavelength. Each simulation domain encompassed the entire head, resulting in a patient-specific grid size. The sonication protocol included a fundamental frequency of 500 kHz, 5% duty cycle, pulse repetition frequency of 100 Hz, and a 30-second on/off cycle for a total duration of 10 minutes [7]. Protocol parameters were stored in YAML format for BabelBrain input. Structural CT and ZTE scans were registered to the corresponding T1w images directly in BabelBrain using Adaptive Stochastic Gradient Descent, and used to generate a bone mask for the simulations (Figure 1). As stated above, in simulations that included a coupling gel pad, an additional 10-mm offset (d_b_) was applied in BabelBrain by (1) increasing the ‘TPO distance’ field by 10 mm and (2) setting the ‘Distance Tx outplane to skin’ field to 10 mm to ensure the added distance was applied outward from the scalp rather than intracranially. Where indicated, simulations included mechanical adjustments to maximize engagement of the intended target. BabelBrain does this by creating a new trajectory that aims at a shifted target, t_2_. However, by “aiming” the beam at t_2_, the maximum acoustic energy is delivered at or near the original target t. As such, we call our true target t the “optimized-target”, and we denote t_2_ as the “nonoptimized-target”. Simulations were run in BabelBrain to compare pressures at both the optimized-target and the nonoptimized-target. We evaluated both targets across simulation platforms because discrepancies between modeling approaches may vary as a function of spatial offset from the focal beam center. Assessing pressure at both locations therefore allows us to determine whether platform-specific differences are uniform near the focus or increase with distance from the region of maximal acoustic energy. BabelBrain simulations report the following metrics: maximum I_SPPA_, spatial-peak temporal-average intensity (I_SPTA_; a diagnostic ultrasound safety metric), and mechanical index within the brain; I_SPPA_ and I_SPTA_ at the target; I_SPPA_ in water; and estimates of temperature and cumulative equivalent minutes (CEM), which is a measure of thermal dose, at the target, brain, skull, and scalp.

### k-Plan Simulations

Simulations were performed using k-Plan version 1.2.0. To ensure that the difference in the results was not due to the registration of the CT / pseudo-CT to the MRI, the T1w images generated from SimNIBS segmentation were imported into k-Plan, along with the registered CT / pseudo-CT volumes generated in BabelBrain (Figure 1). The DPX-500-4CH transducer was selected for all VS simulations, with a grid resolution of six points per wavelength, and the CTX-250-4CH transducer was selected for all Anterior Insula simulations, with a grid resolution of six points per wavelength. Each simulation covered the full head, generating patient-specific grid sizes. As described in the target planning section, we used BabelBrain’s mechanically adjusted trajectory, 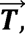 to generate a geometrically matching trajectory, 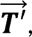 in k-Plan (Figure 1) and combined d_a_ and d_b_ values were entered in the k-Plan interface in the ‘Focal Distance’ field. Free-field I_SPPA_ values were then set to 15, 30, 40, and 60 W/cm². To maintain comparability across platforms, the I_SPPA_ in water reported in BabelBrain’s exported summaries was matched in k-Plan by adjusting the input I_SPPA_ in water values accordingly. For each freefield water ISPPA condition, k-Plan reported maximum and target in situ pressure, I_SPPA_ and I_SPTA_, maximum mechanical index, and change in temperature. However, the k-Plan simulation reports consistently identified the maximum peak pressure outside the brain, often within the transducer itself. Therefore, to enable direct comparison of maximum and target values with BabelBrain outputs, pressure maps exported from k-Plan were used to extract the maximum, optimized-target, and nonoptimized-target in situ I_SPPA_ and I_SPTA_ values. k-Plan additionally reports if the max and target thermal dose exceed 0.1 cumulative equivalent minutes (CEM), but does not report the exact values, and therefore, CEM values cannot be compared against BabelBrain’s output. The extracted outputs were then directly compared with the corresponding BabelBrain simulation results to assess differences in predicted focal intensity and thermal effects between the two software platforms. To quantify discrepancies between the platforms, we calculated the percent difference as: 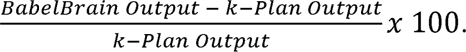

### Statistical Analysis

Linear mixed-effects models were used to assess whether any differences in simulation outputs were due to chance. Simulation outputs (e.g., I_SPPA_, max temperature at the skull) were modeled with a fixed effect of simulation platform. To account for the repeated measures within subjects across the 4 trajectories, we included a random effect of subject.

## Results

Simulation outputs from k-Plan and BabelBrain were compared across four trajectories and two regions for eight subjects using a free-field I_SPPA_ of 60 W/cm², as this value most approximates intensities used in experimental sonication protocols. Importantly, for each comparison, we ensured the exact same trajectory and target location were used between the two platforms as described in the Methods. Substantial discrepancies were observed between k-Plan and BabelBrain simulations for in-situ I_SPPA_ and maximum rise in temperature at the skull at different brain targets and with different fundamental sonication frequencies (Figure 3, Supplementary Figure S5).

**Figure 3.**
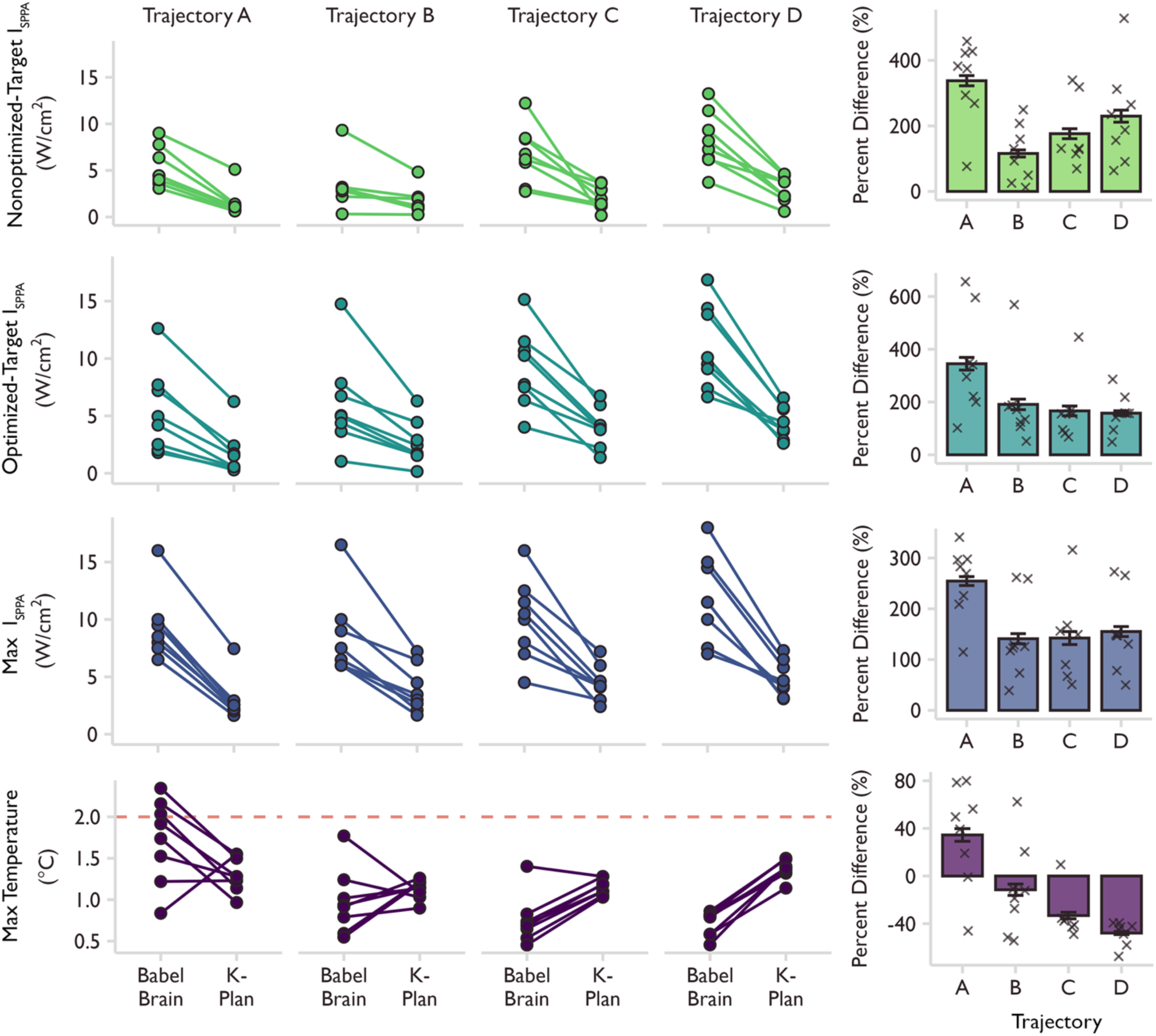
k-Plan and BabelBrain simulation outputs varied across all trajectories. Left panels: Simulation results (n=8) from the four trajectories (A-D) were compared between k-Plan and BabelBrain for the nonoptimized-target, optimized-target, and max derated brain I_SPPA_, and the max change in skull temperature (℃). The nonoptimized-target did not involve any mechanical adjustment of the trajectory to correct for skull-induced aberrations. The optimized-target reflects a mechanically adjusted trajectory to ensure the center of the sonication beam is close to intended target. The max value is the maximum I_SPPA_ in the brain. The location of the max value could differ between simulation platforms, while location of the target values was the same. The dashed red line indicates the max change in temperature safety threshold of 2℃. All trajectories were simulated at 60 W/cm^2^ I_SPPA_ in water. Right panels: Percent differences between BabelBrain and k-Plan outputs are shown for each trajectory across all eight subjects, with mean ± SEM overlaid. One patient was excluded from the nonoptimized-target, Trajectory C bar plot, as they were an outlier (percent difference between k-Plan and BabelBrain was >7000%). Percent difference was calculated as: 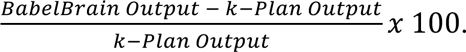

### Difference in focal intensity between simulation platforms

For the four trajectories targeting the VS, we first compared I_SPPA_ values at the nonoptimized-target, as k-Plan does not have an option for automated mechanical adjustments. On average, BabelBrain reported a mean I_SPPA_ value of 5.9+/-3.18 W/cm^2^, while k-Plan reported a mean of 2.05 +/- 1.44 W/cm^2^. We statistically compared these values by fitting a linear mixed-effects model with a fixed effect of simulation platform and a random effect of subject to predict the nonoptimized-target I_SPPA_ value. We found a significant difference between the two simulation platforms (Std. Beta_k-Plan>BabelBrain_ = -1.23, 95% CI:[-1.60, -0.87], n_obs_ = 64, n_subjects_ = 8; Table 1, Figure 3). However, the nonoptimized-target is usually off the center of the sonication beam due to skull-induced aberrations, and it is possible that the simulation platforms may have better alignment in sonication pressures near the center of the sonication beam, where the max pressure is expected, compared to the edge of the sonication beam. Bablebrain offers a ‘mechanical adjustment’ feature that corrects the beam trajectory to ensure the sonication beam is centered around the intended target. Therefore, we next compared the optimized-trajectory pressures at the optimized-target between the two platforms – Note: the target location itself is not different between optimized- and nonoptimized-target, but the sonication beam locations are different; thus, the relative location of the target to sonication beam is different between optimized- and non-optimized-target. BabelBrain reported a mean I_SPPA_ of 7.89 +/- 4.22 W/cm^2^ while k-Plan reported an average of 3.2 +/- 2.02 W/cm^2^. A linear-mixed effects analysis again found a significant effect of simulation platform (Std. Beta_k-Plan>BabelBrain_ = -1.16, 95% CI:[-1.49, -0.83], n_obs_ = 64, n_subjects_ = 8; Table 1, Figure 3). We next tested whether the discrepancy between platforms is due to differences in beam alignment. In such case, we would expect the maximum in situ I_SPPA_ values to be similar while the target values would differ. However, the maximum I_SPPA_ values still showed substantial and significant differences between the simulation platforms (Mean+/-SD_BabelBrain_ : 9.94+/- 3.54 W/cm^2^; Mean+/-SD_k-Plan_: 4.03+/- 1.82 W/cm^2^; Std. Beta_k-Plan>BabelBrain_ = -1.45, 95% CI:[-1.69, -1.2], n_obs_ = 64, n_subjects_ = 8; Table 1, Figure 3) suggesting the differences observed are not simply due to misaligned simulation beams. Additionally, we compared the sonication maps for Subject 1 using Trajectory D (Figure 4). The intensity maps normalized to each simulation peak intensity showed a high degree of overlap (Figure 4b,d,f). However, the absolute intensities differed systematically across the entire sonication focus (Figure 4a,c,e), indicating that differences in sonication results were not simply due to misalignment. Optimized-target I_SPTA_ values also differed between the simulation platforms (Supplemental Figure 2).

**Figure 4.**
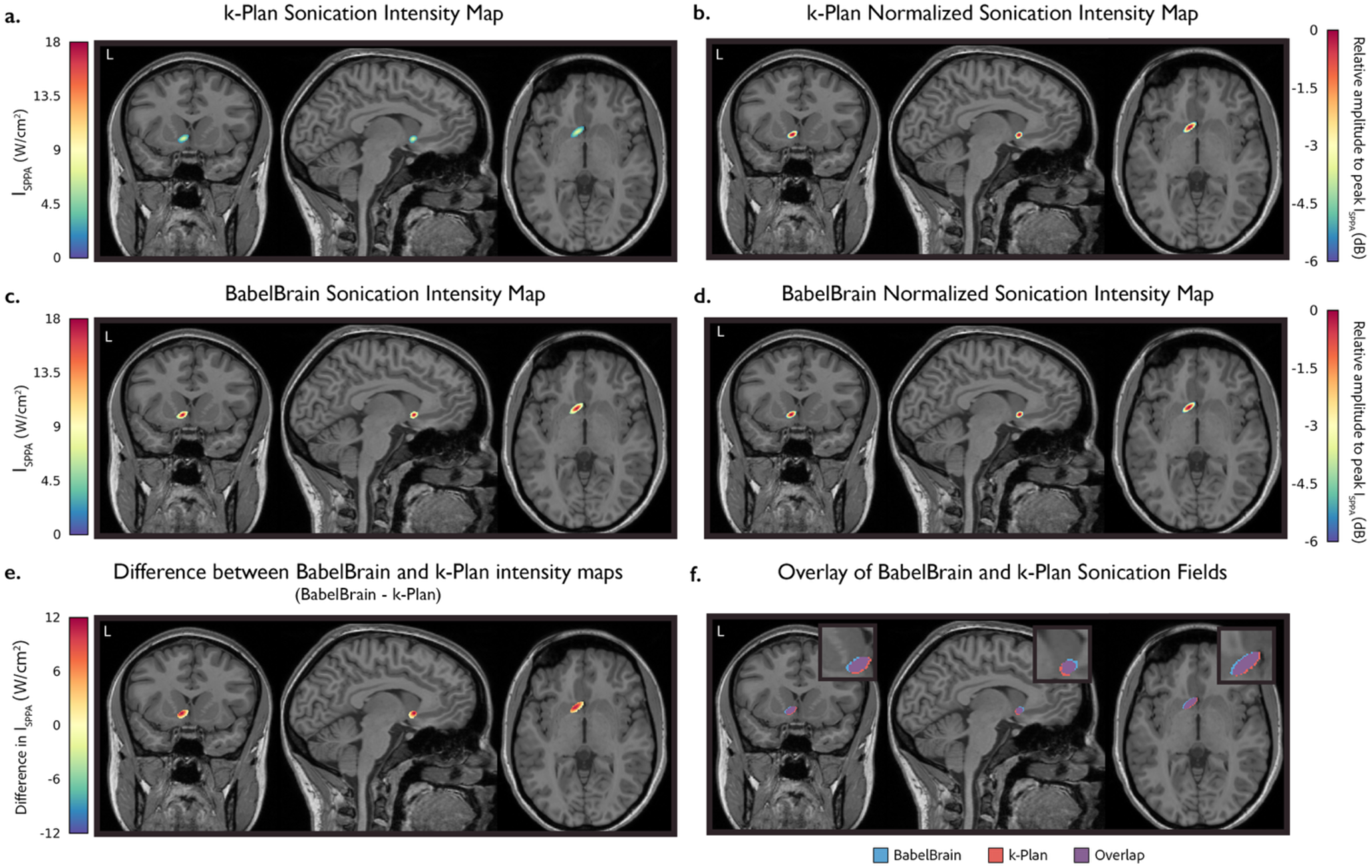
Sonication beam shape and intensity comparison across platforms. **(a–b)** k-Plan sonication intensity maps overlaid on the subject’s T1-weighted MRI with (**a)** showing absolute spatial-peak pulse-average intensity (I_SPPA_; W/cm²), and **(b)** showing normalized intensity to the peak I_SPPA_, expressed in decibels (dB). Both maps are thresholded at −6 dB (25% of peak intensity). **(c–d)** Same as panels **(a–b**), but for the BabelBrain sonication intensity maps. (**e)** Difference between the BabelBrain and k-Plan absolute intensity maps (BabelBrain − k-Plan). Positive vs. negative values indicate regions where BabelBrain predicts higher vs. lower intensities than k-Plan, respectively. (**f)** Spatial overlap of the −6 dB sonication regions from BabelBrain and k-Plan. Insets show enlarged views of the sonication focus, near the target.

**Table 1.**
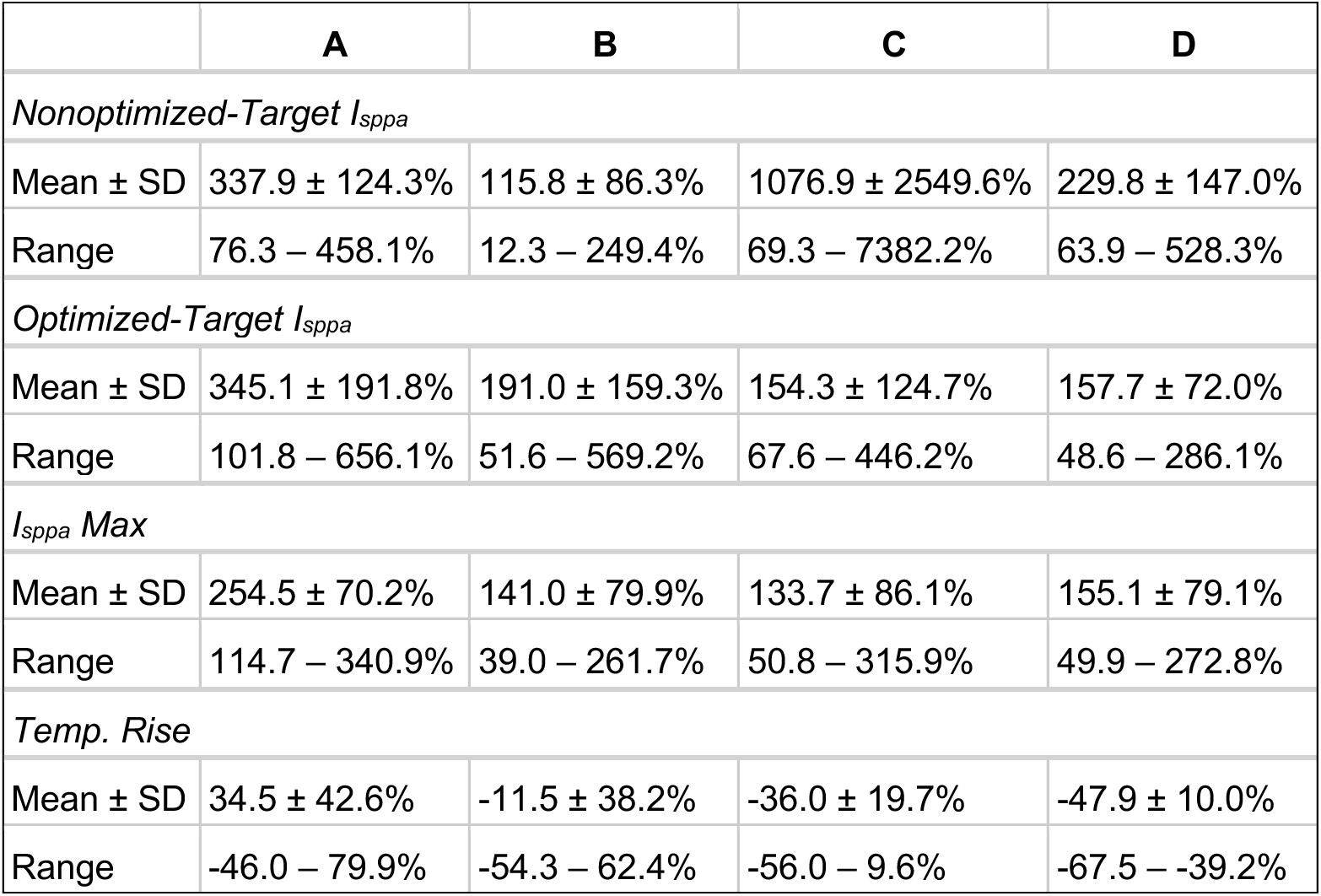
Percent change in key metrics across simulation platforms. Percent change in intensity and temperature rise between simulation platforms across the four trajectories. We compared the intensity values between the simulation platforms at a free-field I_SPPA_ of 60 W/cm², across three different focal points (nonoptimized-target, optimized-target, and max) and estimated max temperature rise in the skull. Mean, standard deviation (SD), and range are reported. Percent difference was calculated as: 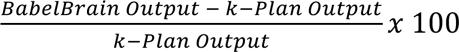

|  | **A** | **B** | **C** | **D** |
| --- | --- | --- | --- | --- |
| *Nonoptimized-Target I<sub>sppa</sub>* |  |  |  |  |
| Mean ± SD | 337.9 ± 124.3% | 115.8 ± 86.3% | 1076.9 ± 2549.6% | 229.8 ± 147.0% |
| Range | 76.3 – 458.1% | 12.3 – 249.4% | 69.3 – 7382.2% | 63.9 – 528.3% |
| *Optimized-Target I<sub>sppa</sub>* |  |  |  |  |
| Mean ± SD | 345.1 ± 191.8% | 191.0 ± 159.3% | 154.3 ± 124.7% | 157.7 ± 72.0% |
| Range | 101.8 – 656.1% | 51.6 – 569.2% | 67.6 – 446.2% | 48.6 – 286.1% |
| *I<sub>sppa</sub> Max* |  |  |  |  |
| Mean ± SD | 254.5 ± 70.2% | 141.0 ± 79.9% | 133.7 ± 86.1% | 155.1 ± 79.1% |
| Range | 114.7 – 340.9% | 39.0 – 261.7% | 50.8 – 315.9% | 49.9 – 272.8% |
| *Temp. Rise* |  |  |  |  |
| Mean ± SD | 34.5 ± 42.6% | -11.5 ± 38.2% | -36.0 ± 19.7% | -47.9 ± 10.0% |
| Range | -46.0 – 79.9% | -54.3 – 62.4% | -56.0 – 9.6% | -67.5 – -39.2% |

Given that focal intensity scales linearly with stimulation intensity, we also tested whether the magnitude of the discrepancy between the two platforms would remain constant across different stimulation intensities. Free-field I_SPPA_ in water was simulated at 15, 30, 45, and 60 W/cm² in both platforms using trajectory D. As expected, across all conditions, a systematic, constant-magnitude difference was observed between the two software packages (Supplemental Figure 3).

### Difference in thermal rise between simulation platforms

In addition to testing for differences in the focal intensities, we compared the expected maximum temperature rise at the skull, which is generally much higher than the brain temperature rise, between the two platforms. As with the intensities, we found that platform identity predicted different temperature rises (Std. Beta_k-Plan>BabelBrain_ = 0.49, 95% CI:[0.001, 0.98], n_obs_ = 64, n_subjects_ = 8; Table 1, Figure 3). However, simulated changes in maximum temperature rise showed less systematic differences between platforms. Indeed, while the max I_SPPA_ values between k-Plan and BabelBrain were highly correlated with each other (r = 0.89), changes in temperature were much less so (r = .15; Supplemental Figure 4). The low correlation between the simulated temperature changes may be driven by BabelBrain’s wider range of estimated temperature changes across the 4 trajectories (SD_BabelBrain_ = 0.54), while k-Plan estimated a more stable rise in temperature across the different trajectories (SD_k-Plan_ = 0.17; Figure 3).

We also assessed whether the observed differences in thermal rise and focal intensity were specific to the ventral striatum or to the 500 kHz fundamental frequency. We found similarly discrepant results when targeting the ventral Anterior Insula using a transducer with a fundamental frequency of 250kHz (Supplemental Figure 5; Supplemental Table 1). Taken together, these findings indicate that despite identical input parameters and targeting, simulation outputs differed markedly between k-Plan and BabelBrain across brain areas, trajectories, and sonication frequencies.

### Differences between simulation platforms when decreasing effective aperture

Recent calls to standardize reporting of ultrasound sonications highlight the need to report the “effective aperture” of the transducer [23]. The effective aperture of the ultrasound beam can differ from the nominal aperture of the transducer if a coupling medium adds to the distance between the transducer and scalp, affecting the ultrasound beam propagation. For example, adding a coupling medium, such as a 10 mm-thick gel pad, can reduce the effective aperture below the nominal aperture, potentially altering simulated safety metrics. To quantify such effects across platforms, we simulated adding a 10 mm gel pad between the transducer and scalp for one of the orthogonal trajectories in five of the subjects (Figure 5). The results reinforced the overall trends observed in Figure 3 while revealing additional effects. For acoustic intensity measures, optimized-target I_SPPA_ and max I_SPPA_ values remained consistently lower in k-Plan compared to BabelBrain, with minimal changes when accounting for the gel pad distance. However, both platforms reported an increase in maximum temperature change when the 10 mm gel pad was included, as would be expected with higher beam density in the skull with lower effective aperture. However, k-Plan demonstrated greater magnitude difference than BabelBrain in predicted temperature rise when incorporating the added coupling medium distance. Thus, the addition of a 10 mm scalp-transducer offset resulted in an increase in predicted maximum temperature, likely due to higher ultrasound density per crossed bone area, highlighting the sensitivity of sonication simulations to even minor adjustments in setup parameters, and the potential for such factors to influence dosing estimates relative to safety parameters. Critically, while this analysis compares the simulation platforms’ outputs when adding a coupling medium, these analyses cannot capture distortions related to other material sitting between the transducer and skull, such as air pockets and bubbles, which are frequently introduced by the participants’ hair. As these platforms cannot estimate how ubiquitous these distortions are in any given LIFU session, they represent additional sources of error in the simulation outputs.

**Figure 5.**
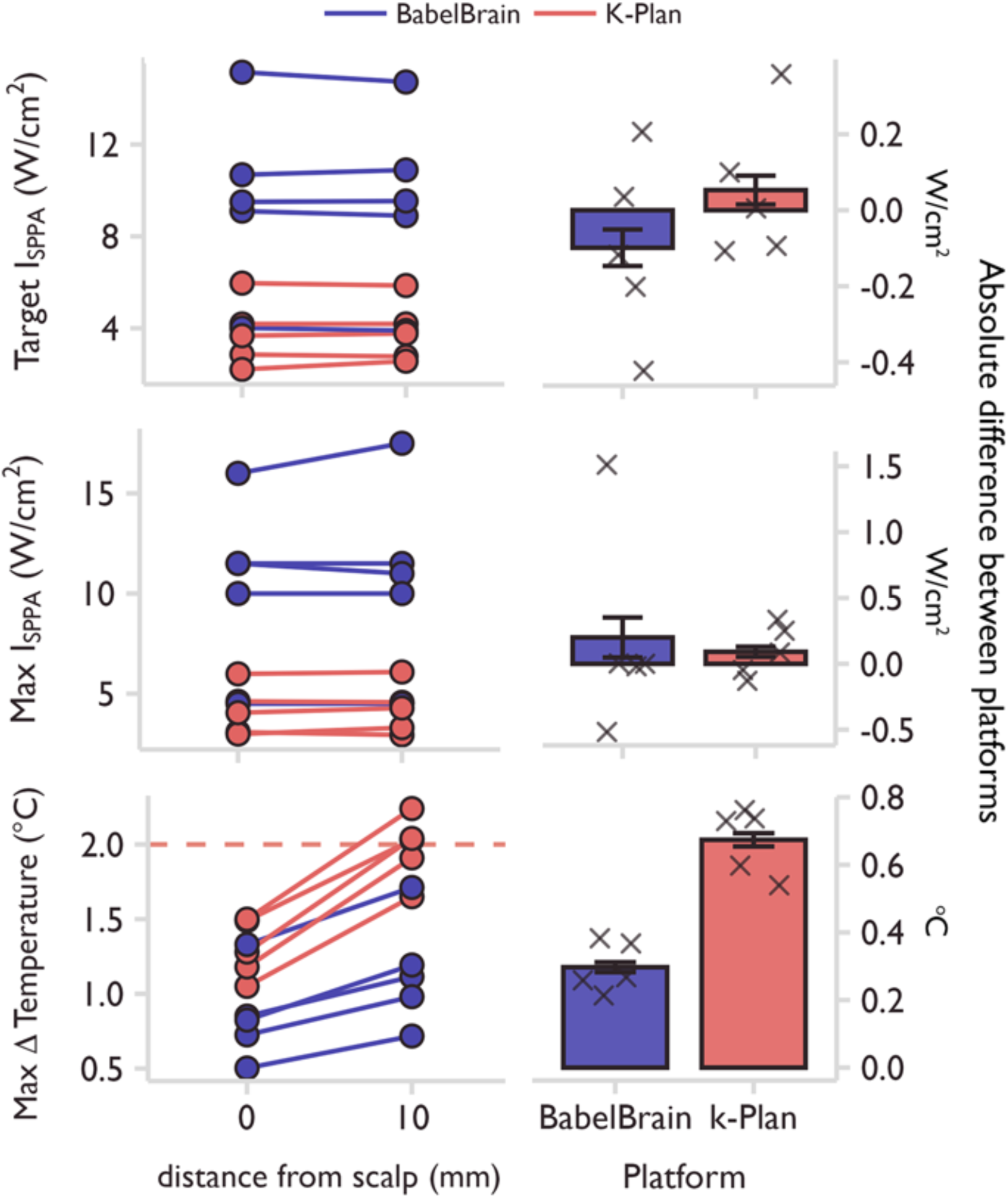
Reducing the effective aperture with a coupling medium is modeled differently between platforms. Left panels: simulation results at two distances from the scalp (0 mm and 10 mm) in five subjects (n=5) were compared to the optimized-target, max derated I_SPPA_ in the brain, and the max change in skull temperature (℃). One of the orthogonal trajectories was simulated in each subject with an additional 10 mm added outside of the scalp. The dashed red lines indicate a temperature rise of 2℃. Both distances were simulated at 60 W/cm^2^ I_SPPA_ in water. Right panels: Absolute differences between 0 mm and 10 mm from the scalp in each platform are shown for each intensity level across all subjects, with mean ± SEM displayed.

Finally, we compared the platforms in terms of computation time. When simulating the four trajectories, computation time in BabelBrain was on average 50.47 +/- 5.26 minutes across subjects. In k-Plan, the computation time was on average 7.1 +/- 0.03 days. Despite the long average computation time, individual simulations in k-Plan could be completed in a few hours depending on server availability.

### Differences in focal intensity and thermal rise simulations between CT and pseudo-CT inputs

CT- and pseudo-CT–based simulations were compared in five subjects using the same predefined VS trajectories to evaluate whether the skull masks produced similar results. Because this comparison does not evaluate the platform itself but rather focuses on the skull masks derived from the CT and pseudo-CT images, we chose to run the simulations in BabelBrain. The pseudo-CT masks were generated from GE ZTE imaging sequences using the built-in BabelBrain algorithm. Percent difference was calculated using the CT simulated result as the reference. As above, we assessed for differences in focus intensity and temperature rise using linear mixed effects models with a random effect of subject. Across subjects and trajectories, simulation outputs showed substantial variability (Figure 6, Supplemental Table 2, Supplemental Figure 6).

**Figure 6.**
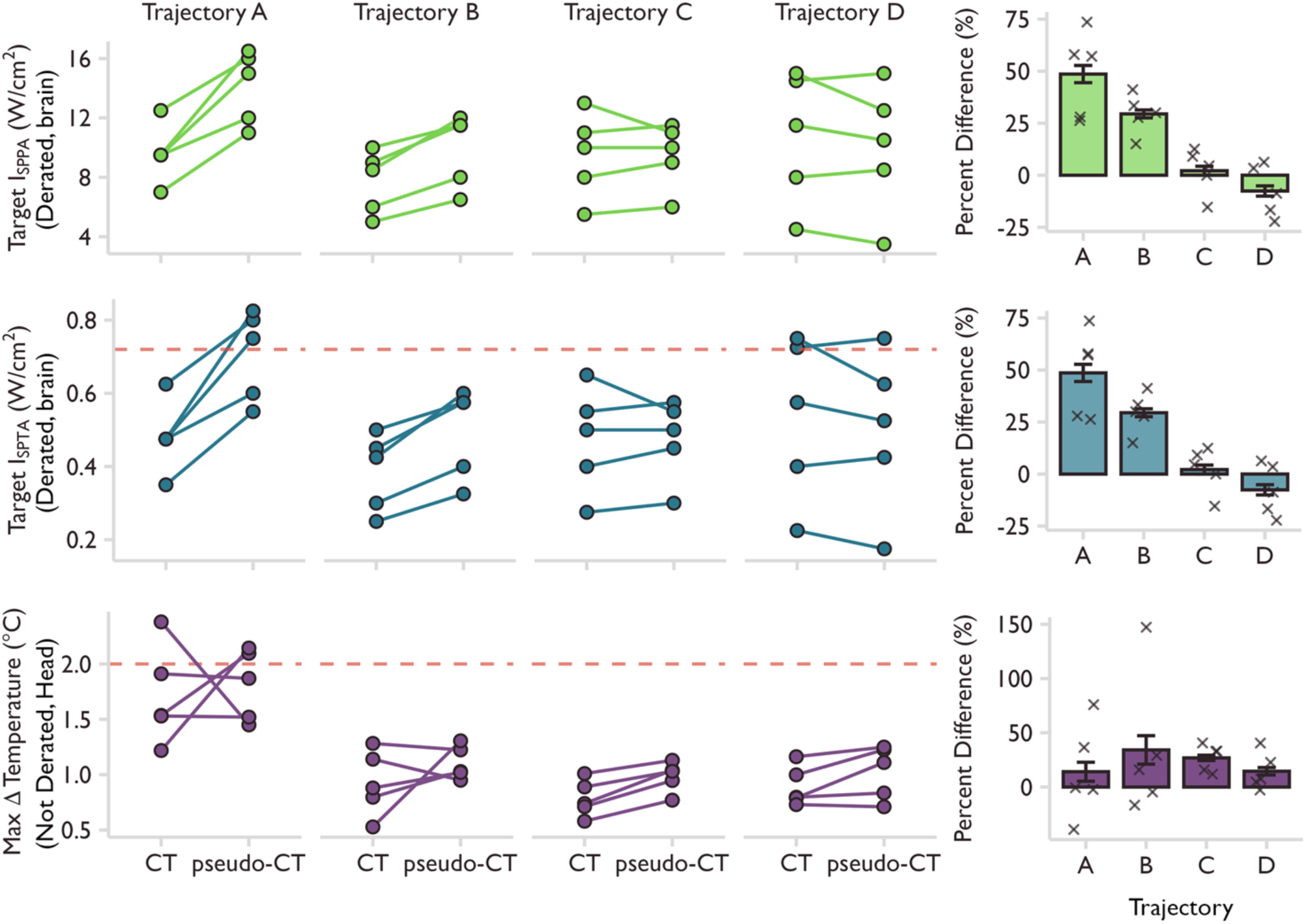
Comparison of CT and pseudo-CT-based simulations. Left panels: Simulation outputs for five subjects, each with both CT and ZTE images, are shown across the four sonication trajectories (A–D). For each subject, simulations were run in BabelBrain using either CT-derived or ZTE-derived (pseudo-CT) skull models. Outputs include the target derated brain I_SPPA_, target derated brain I_SPTA_, and the maximum change in skull temperature (℃). The dashed red lines represent the safety limits of 0.72 W/cm^2^ I_SPTA_ and a 2℃ temperature rise. All simulations were performed at 60 W/cm² I_SPPA_ in water with the transducer positioned 10 mm from the scalp. Right panels: Percent differences between CT and pseudo-CT simulations are shown for each trajectory across all subjects, with mean ± SEM. Percent difference was calculated as: 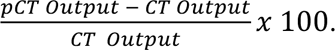

When testing for differences in I_SPPA_ at the optimized-target, we found a trend difference between CT and pseudo-CT (Std. Beta_pCT>CT_ = 0.47, 95% CI:[-0.01, 0.94], n_obs_ = 20, n_subjects_ = 5; Figure 6, Supplemental Table 2). Post-hoc tests revealed that differences were driven by the nonorthogonal trajectories (Trajectory A: t(4) = -5.7, p < .005; Trajectory B: t(4) = -5.97, p < .005), while the orthogonal trajectories resulted in similar intensity values (Trajectory C: t(4) = 0, p > .05; Trajectory D: t(4) = 1.25, p >.05). While there were no significant differences in estimated temperature rise between the pseudo-CT and CT (Std. Beta_pCT>CT_ = 0.35, 95% CI:[-0.29, 0.99], n_obs_ = 20, n_subjects_ = 5; Figure 6, Supplemental Table 2), and between simulated intensities in orthogonal trajectories, there was still meaningful variation in thermal rise and sonication intensities for some subjects. For example, in Trajectory A, two subjects had estimated temperature rises above the 2°C safety threshold when using the pseudo-CT as the planning image but not the CT; another subject showed the reverse pattern. As with the intensity results, with the more orthogonal trajectories, the direction of the discrepancy between platforms ranging up to 20% was inconsistent, with pseudo-CT simulations sometimes yielding higher values and other times lower ones in different metrics (e.g., I_SPPA_). This finding indicates that pseudo-CT-based simulations do not reliably produce more conservative estimates. Furthermore, we compared the sonication maps for one subject using Trajectory D (Supplemental Figure 7). The intensity maps normalized to each simulation peak intensity showed a high degree of overlap (Supplemental Figure 7b,d, f). The absolute intensities differed systematically along the beam path (Supplemental Figure 7a,c,e), indicating that identified differences are not due to beam misalignment. Together, these results highlight the need for further optimization of pseudo-CT imaging sequences and reconstruction algorithms before they can be considered a reliable substitute for CT-based skull modeling in LIFU simulations.

## Discussion

While individually simulating LIFU sonication is an important step for ensuring safety and accuracy, our results demonstrate substantial discrepancies in key parameters across the two commonly used simulation platforms (BabelBrain and k-Plan), and between CT-based and pseudo-CT-based simulations, even when identical input parameters and trajectories were used. This is especially relevant for safety and reproducibility. We found that across two targets, two sonication frequencies, four representative trajectories, and eight subjects, I_SPPA_ and estimated temperature rise varied across the platforms. BabelBrain estimated consistently higher focal intensities compared to k-Plan, but estimated overall lower temperature rises. However, the platform that estimated higher thermal rises differed by trajectory and showed weaker correlations between platforms compared to focal intensities (r_temp_ = 0.15; r_max-ISPPA_ = 0.89). These changes were consistent across different stimulation intensities but varied further when accounting for the distance between the scalp and the transducer. Finally, we also found differences in key simulated outputs when comparing CT versus pseudo-CT scans as inputs for skull modeling, though these differences were smaller for the more orthogonal trajectories.

### Disagreement between platforms across key safety & dosing metrics

Across all trajectories, k-Plan estimated lower focal intensities compared to BabelBrain, with BabelBrain frequently estimating double or triple the intensity estimated by k-Plan. This is clinically meaningful when interpreting both positive and negative results from the existing LIFU literature, and with regard to safety guidelines proposed for LIFU. For example, using a 60 W/cm² ISPPA in-water input in our simulations, a temperature rise exceeding the 2 °C threshold recommended by the ITRUSST consensus guidelines [24] was observed in three subjects for one platform but not the other. This effect could be further exacerbated at higher sonication pressures than the 60 W/cm² used in our simulations. Additionally, while k-Plan estimated higher temperature rises for the orthogonal trajectories (C and D), within trajectory A, BabelBrain predicted three subjects would experience a temperature rise exceeding the 2 °C guideline recommended by the ITRUSST consensus guidelines [24]. Interestingly, the transducer was placed near the temple for this trajectory, corresponding to a region of the skull with more heterogeneous bone architecture; this was the only trajectory where BabelBrain estimated consistently higher temperature rises compared to k-Plan. These disagreeing safety-related outputs, even within our small sample size, highlight the risk of relying solely on a single simulation platform without using reasonable safety margins.

To further approximate real-world sonication conditions, we simulated the effect of a 10 mm gel pad placed between the transducer and scalp. While BabelBrain and k-Plan generally showed minimal changes in optimized-target I_SPPA_ and max I_SPPA_ values, both platforms predicted significant increases in temperature rise with the added distance (Figure 5). Notably, in three subjects k-Plan simulations reached the 2 °C safety threshold with the gel pad included. These results underscore that even modest increases in transducer-scalp distance can substantially alter predicted temperature outcomes. Moreover, the effect was not consistent across platforms, complicating safety interpretation.

It is critical to note that neither overestimation nor underestimation of simulation outputs translate to safer sonications. Underestimation of simulated acoustic pressures can be particularly problematic when studies aim to achieve a specific in situ pressure at the target across participants, as it may prompt investigators to increase delivered sonication amplitudes to reach the desired (apparently sub-threshold) pressure, potentially resulting in true acoustic pressures that exceed safety guidelines. Alternatively, overestimation of temperature rise may lead researchers to select overly cautious parameters that underdose the target, increasing the risk of null effects and delaying scientific progress. Together, these methodological differences highlight the need for standardized, validated pipelines for deriving skull acoustic and thermal properties to ensure consistent, effective, and safe dosing.

### Comparison between simulation approaches

Critically, it remains unclear which platform provides estimates that are closer to the ground truth. Determining this would require validation against hydrophone-based measurements of sonication pressures and measurements of temperature rise. In the absence of these tests, we highlight the differences in simulation approach and hypothesize that the core differences between platforms are primarily driven by differences in estimating the bone acoustic properties from the CT (or pseudo-CT).

BabelBrain and k-Plan use different computational models of acoustic wave propagation in their simulations: BabelBrain uses BabelViscoFDTD, while k-Plan uses k-Wave. These models were recently compared and found to have similar outputs, when controlling for the inputs (such as the acoustic properties of the skull) – at target, differences between k-Wave and BabelViscoFDTD in focal pressure did not exceed 2.7% across different benchmarks [25]. This suggests that the underlying acoustic wave propagation models are not driving the differences presented here and, instead, stem from inputs to these models. Such inputs include: (1) medium parameters, including acoustic properties for the skull and soft-tissue, 2) transducer characteristics, and 3) other numerical parameters for the models, such as grid resolution. As identical values were used for the transducer characteristics and each model was run with a grid resolution of 6 points per wavelength, this suggests the differences are driven by how each platform models the acoustic properties of skull and soft-tissue, which the two platforms handle quite differently (Table 2).

**Table 2.** Estimation of acoustic properties of the skull in k-Plan and BabelBrain.

| Modeling Step | Description | k-Plan | BabelBrain |
| --- | --- | --- | --- |
| Segmentation approach | Each platform segments the planning image (either a CT or pseudo-CT) into bone, soft tissue, and background | k-Plan segments the image using a density-based threshold:<br>bone: $\geq 1150 \text{ kg/m}^3$<br>soft tissue: $\geq 900 \text{ kg/m}^3$<br>background: $< 900 \text{ kg/m}^3$ | BabelBrain uses a multi-step approach, including taking as inputs skin, skull, and brain masks derived from the SimNIBS charm processing tool, which is based on T1/T2 images. These masks are further refined by coregistering the skull mask to the planning image and thresholding it at $\geq 300 \text{ HU}$ |
| Density mapping | The planning images can be converted from HU to density ($\text{kg/m}^3$) before conversion to speed of sound | k-Plan uses a calibration file, either a default file, which is an average across vendors, or a user-specified, scanner-specific calibration file obtained by the user scanning a phantom.<br>For thermal simulations, k-Plan uses a constant density of $1850 \text{ kg/m}^3$. | BabelBrain converts from HU to the speed of sound directly, without mapping to density values first (see below). |
| Sound speed mapping | Either the density values or HU from the previous steps are converted to the estimated speed of sound through the skull. | k-Plan applies a linear formula to convert from density to speed of sound:<br>$1.33 \times \text{density} + 166.7$ (from Marquet et al. 2009) | BabelBrain follows the method in Webb-Marsac et al., which converts directly from HU to sound speed using scanner-specific fitting parameters that account for scanner model, energy level, and reconstruction kernel. |
| Attenuation mapping | The attenuation rate determines how much energy the ultrasound wave loses as it passes through each voxel. | k-Plan uses a uniform attenuation coefficient of $13.3 \text{ dB}/(\text{cm.MHz}^y)$ with a power law exponent ($y$) of 1, applied identically to each voxel in the bone mask. | BabelBrain uses HU values and scanner-specific fitting parameters to estimate the rate of attenuation for each voxel in the bone mask. |
| Soft tissue & background properties | Fixed values are used to estimate the sound speed and attenuation for soft tissue and background voxels | k-Plan uses:<br>*soft tissue*<br>density: $1045 \text{ kg/m}^3$<br>sound speed: 1550<br>attenuation: $0.59 \text{ dB}/(\text{cm.MHz}^y)$, $y=1.2$<br>*background*<br>density: $996 \text{ kg/m}^3$<br>sound speed: 1509<br>attenuation: $2.17\text{e-}3 \text{ dB}/(\text{cm.MHz}^y)$, $y=2$ | BabelBrain uses:<br>*soft tissue*<br>density: $1041 \text{ kg/m}^3$<br>sound speed: 1562<br>attenuation: $0.59 \text{ dB}/(\text{cm.MHz}^y)$, $y=1.2$<br>*background*<br>density: $1000 \text{ kg/m}^3$<br>sound speed: 1500<br>attenuation: 0 |
| Absorption-to-heat conversion | Each platform uses the attenuation coefficient for thermal modeling. | k-Plan uses the full attenuation coefficient when converting to heat. | BabelBrain assumes that the fraction of skull attenuation that is converted to heat is 0.16, based on Pinton et al. 2011 |

Deriving individualized bone acoustic properties (e.g., attenuation coefficient) from CTs (or pseudo-CTs) is not standardized [26, 27]. Briefly, this process requires segmenting the planning image (CT or pseudo-CT) into bone, soft tissue, and background, converting the Hounsfield Units (HU) to attenuation coefficients, and modeling the conversion of sound wave absorption to heat. While the two platforms handle each step slightly differently (Table 2), we highlight three steps that particularly diverge. First, k-Plan converts from HU to the speed of sound at each voxel by first converting to density and then speed of sound, using either a user-specified or general scanner calibration file. BabelBrain converts from HU to speed of sound directly using scanner-vendor specific parameters derived from Marsac et al. 2017 [28]. Differences in the speed of sound values will change the timing of the waves and will thus change the expected phase when the beam exits the skull. Such difference is highly relevant when using transducers with multi-phase arrays that can incorporate phase correction and may result in even larger discrepancies between the simulation platforms. However, modeling assumptions related to phase correction are not applicable in this analysis as neither the DPX-500, 4-element device, nor the CTX-250, 4-element device has phase-refocusing capabilities under normal operation. Second, k-Plan applies a uniform attenuation coefficient of 3.3 dB/(cm.MHz^y^) with a power law exponent of 1, applied identically to each voxel in the bone mask, while BabelBrain estimates the attenuation coefficient for each voxel, following the procedure in Marsac et al. 2017 [28]. The constant high attenuation coefficient used by k-Plan may produce the observed more conservative acoustic outputs, generally resulting in lower predicted *in situ* intensities compared with BabelBrain. Third, when estimating heat absorption, k-Plan uses the full attenuation coefficient, while BabelBrain assumes only 16% of the attenuation coefficient will be converted to heat. This difference likely accounts for the consistently higher temperature rises reported by k-Plan and reflects a more conservative thermal model compared with BabelBrain’s anatomy-specific approach.

While previous papers that compared the underlying acoustic wave models suggested that different simulation approaches are roughly equivalent [25], our work shows that differences in how these platforms estimate the acoustic properties from the skull matters significantly when trying to safely and equivalently dose patients. The hypothesis is also supported by a recent paper, which compared the accuracy of k-Wave simulations through different human skulls [27], which reported that skulls introduced errors of around 35% in pressure amplitude and transmission loss, with even larger errors for individual skulls, highlighting the need for increasing the accuracy of skull attenuation coefficient mapping to reduce these errors [27]. Our results generally support these findings, where the simulated beam paths largely overlap but the peak in situ intensities show consistent differences between k-Plan and BabelBrain.

### Disagreement between CT & pseudo-CT planning images across key safety & dosing metrics

Finally, comparisons between ZTE- and CT-based simulations also revealed notable variability. For example, in one subject, trajectory A exceeded the 2 °C safety guideline in the CT-based simulation, whereas the ZTE-based result did not. Conversely, in another subject, trajectory D surpassed the 0.72 W/cm² I_SPTA_ threshold in the CT-based simulation but not in the ZTE-based simulation. These examples illustrate that ZTE-derived pseudo-CTs cannot be assumed to provide more conservative safety estimates. Although pseudo-CT simulations showed smaller discrepancies to CT-based simulations for orthogonal trajectories, the individual-level differences, up to 20% for pressure and 40% for temperature, are large enough to be problematic when estimates are near safety thresholds, as such deviations can determine whether a trajectory is deemed safe or unsafe.

### Limitations

This study should be interpreted in light of some important limitations. First, in the absence of ground-truth measurements, we cannot determine which simulation platform is more accurate. Establishing such validation benchmarks represents an important next step for the field. Second, the small sample size resulted in wide confidence intervals around the precise magnitude and direction of the observed differences. Nevertheless, the large differences detected even within this limited cohort suggest that discrepancies between the simulation platforms are systemic and unlikely to be rare observations. Third, we used the default CT calibration file in our k-Plan simulations. If our scanners’ measurements significantly differ from the default calibration values, this may reduce the accuracy of the k-Plan estimates of focal intensity. However, k-Plan does not use this calibration file for its thermal simulations. As such, differences in scanner calibration do not affect the estimated differences in temperature rise.

Individualized simulations of LIFU protocols, as compared to assuming a one-size-fits-all approach across participants with varying head sizes and skull thicknesses, remains a critical step towards realizing the power of LIFU neuromodulation as a new treatment for neurological and psychiatric conditions. While the field has already come a long way, our findings highlight the critical importance of transparent reporting of simulation parameters and inputs [23], as well as conducting simulations that accurately reflect the specific details of the experimental setup. Such practices are essential for ensuring both the safety and reproducibility of LIFU studies. Without standardized definitions, reference measurements, and systematic validation against hydrophone and thermometry data, it remains uncertain which simulation platform more faithfully represents the true *in situ* sonication dose. Establishing consensus standards and validating simulation tools against ground-truth measurements will be necessary steps for advancing LIFU toward reliable and clinically safe applications.

## Data Availability

All data produced in the present study are available upon reasonable request to the authors.

## Acknowledgments

KM is funded by the Weill Neurohub and DA060431.

## References

1. Murphy, K.R., et al., A practical guide to transcranial ultrasonic stimulation from the IFCN-endorsed ITRUSST consortium. Clin Neurophysiol, 2025. 171: p. 192–226.

2. Keihani, A., et al., Transcranial Focused Ultrasound Neuromodulation in Psychiatry: Main Characteristics, Current Evidence, and Future Directions. Brain Sci, 2024. 14(11).

3. Bowary, P. and B.D. Greenberg, Noninvasive Focused Ultrasound for Neuromodulation: A Review. Psychiatr Clin North Am, 2018. 41(3): p. 505–514.

4. Legon, W., et al., Noninvasive neuromodulation of subregions of the human insula differentially affect pain processing and heart-rate variability: a within-subjects pseudo-randomized trial. Pain, 2024.

5. Strohman, A., et al., Low-Intensity Focused Ultrasound to the Human Dorsal Anterior Cingulate Attenuates Acute Pain Perception and Autonomic Responses. J Neurosci, 2024. 44(8).

6. Yaakub, S.N., et al., Non-invasive ultrasonic neuromodulation of the human nucleus accumbens impacts reward sensitivity. Nat Commun, 2025. 16(1): p. 10192.

7. Cain, J.A., et al., Real time and delayed effects of subcortical low intensity focused ultrasound. Sci Rep, 2021. 11(1): p. 6100.

8. Riis, T.S., et al., Noninvasive Modulation of the Subcallosal Cingulate and Depression With Focused Ultrasonic Waves. Biol Psychiatry, 2024.

9. Riis, T.S., et al., Noninvasive targeted modulation of pain circuits with focused ultrasonic waves. Pain, 2024. 165(12): p. 2829–2839.

10. Riis, T.S., et al., Noninvasive modulation of essential tremor with focused ultrasonic waves. J Neural Eng, 2024. 21(1).

11. Rezai, A., et al., Focused Ultrasound Neuromodulation: Exploring a Novel Treatment for Severe Opioid Use Disorder. Biol Psychiatry, 2025. 98(1): p. 56–64.

12. Rezai, A., et al., Brain injury during focused ultrasound neuromodulation for substance use disorder. Brain Stimul, 2025. 18(6): p. 2050–2053.

13. Aubry, J.F., et al., ITRUSST consensus on biophysical safety for transcranial ultrasound stimulation. Brain Stimul, 2025. 18(6): p. 1896–1905.

14. Leung, S.A., et al., A rapid beam simulation framework for transcranial focused ultrasound. Sci Rep, 2019. 9(1): p. 7965.

15. Miscouridou, M., et al., Classical and Learned MR to Pseudo-CT Mappings for Accurate Transcranial Ultrasound Simulation. IEEE Trans Ultrason Ferroelectr Freq Control, 2022. 69(10): p. 2896–2905.

16. BrainBox. k-Plan Ultrasound Planning (Version 1.1.4). 2025; Available from: https://brainbox-neuro.com/products/k-plan.

17. Treeby, B.E. and B.T. Cox, k-Wave: MATLAB toolbox for the simulation and reconstruction of photoacoustic wave fields. J Biomed Opt, 2010. 15(2): p. 021314.

18. Pichardo, S., BabelBrain: An Open-Source Application for Prospective Modeling of Transcranial Focused Ultrasound for Neuromodulation Applications. IEEE Trans Ultrason Ferroelectr Freq Control, 2023. 70(7): p. 587–599.

19. Shen, J. Tools for NIfTI and ANALYZE image. 2025; Available from: https://www.mathworks.com/matlabcentral/fileexchange/8797-tools-for-nifti-and-analyze-image.

20. Puonti, O., et al., Accurate and robust whole-head segmentation from magnetic resonance images for individualized head modeling. Neuroimage, 2020. 219: p. 117044.

21. Fairbanks, T., et al., Pipeline for Planning and Execution of Transcranial Ultrasound Neuromodulation Experiments in Humans. J Vis Exp, 2024(208).

22. Kim, C., M. Eames, and D.G. Paeng, Improving Sonication Efficiency in Transcranial MR-Guided Focused Ultrasound Treatment: A Patient-Data Simulation Study. Bioengineering (Basel), 2023. 11(1).

23. Martin, E., et al., ITRUSST consensus on standardised reporting for transcranial ultrasound stimulation. Brain Stimul, 2024. 17(3): p. 607–615.

24. Aubry, J.-F.A., D; Schafer, M; Fouragnan, M; Charles Caskey, Robert Chen, Ghazaleh Darmani, Ellen J. Bubrick, Jérôme Sallet, Christopher Butler, Charlotte Stagg, Miriam Klein-Flügge, Seung-Schik Yoo, Brad Treeby, Lennart Verhagen; Butts Pauly, Kim ITRUSST Consensus on Biophysical Safety for Transcranial Ultrasonic Stimulation. arXiv, 2023. arXiv:2311.05359.

25. Aubry, J.F., et al., Benchmark problems for transcranial ultrasound simulation: Intercomparison of compressional wave models. J Acoust Soc Am, 2022. 152(2): p. 1003.

26. Angla, C., et al., Transcranial ultrasound simulations: A review. Med Phys, 2023. 50(2): p. 1051–1072.

27. Krokhmal, A., et al., A comparative study of experimental and simulated ultrasound beam propagation through cranial bones. Phys Med Biol, 2025. 70(2).

28. Marsac, L., et al., Ex vivo optimisation of a heterogeneous speed of sound model of the human skull for non-invasive transcranial focused ultrasound at 1 MHz. Int J Hyperthermia, 2017. 33(6): p. 635–645.

