## Supplement for "Empirical limitations of current low-intensity focused ultrasound simulation platforms"

**Supplemental Figures:**

- Figures S1-S7
- Tables S1-S2

**Figure S1.**

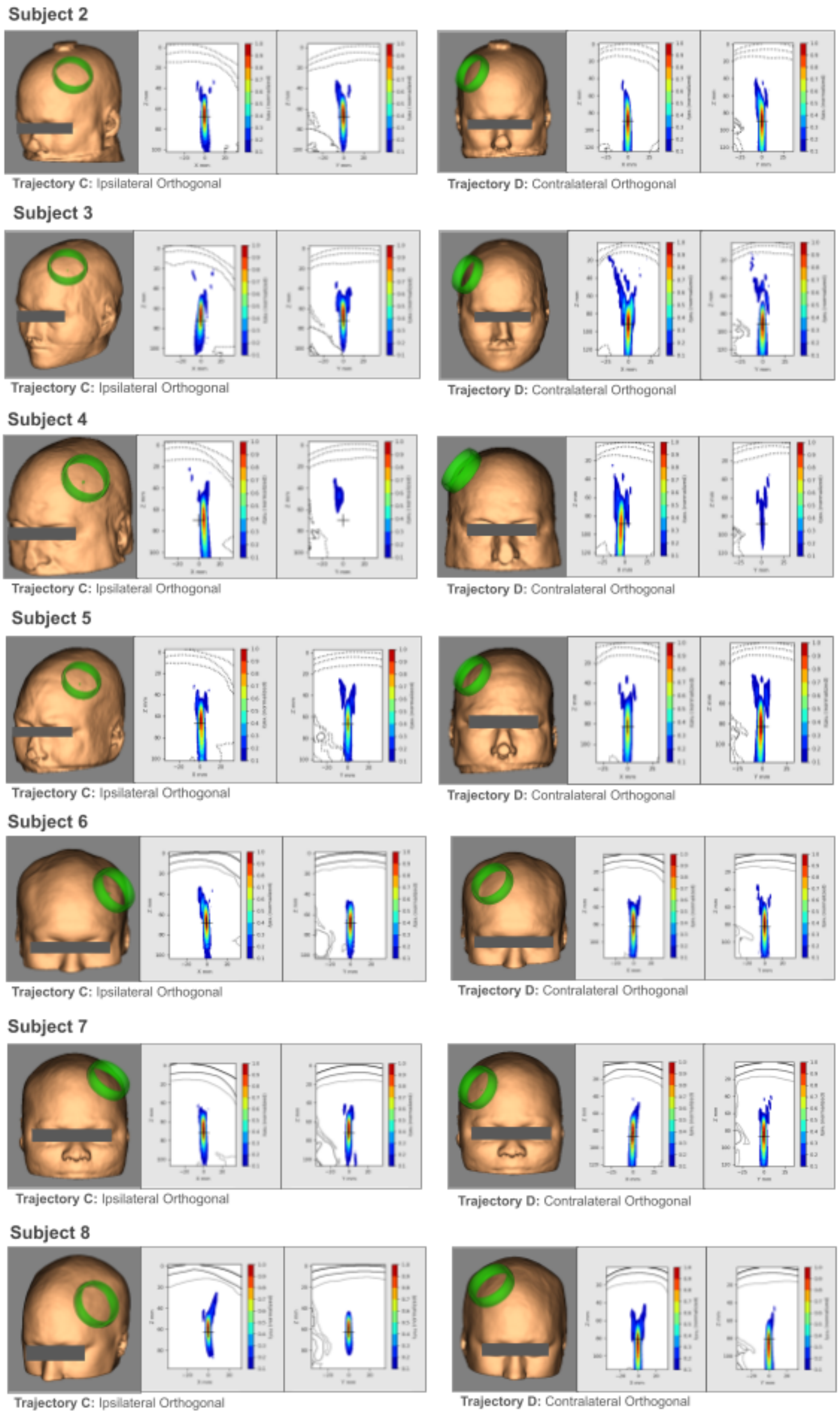
Subject 2
Trajectory C: Ipsilateral Orthogonal
Trajectory D: Contralateral Orthogonal
Subject 3
Trajectory C: Ipsilateral Orthogonal
Trajectory D: Contralateral Orthogonal
Subject 4
Trajectory C: Ipsilateral Orthogonal
Trajectory D: Contralateral Orthogonal
Subject 5
Trajectory C: Ipsilateral Orthogonal
Trajectory D: Contralateral Orthogonal
Subject 6
Trajectory C: Ipsilateral Orthogonal
Trajectory D: Contralateral Orthogonal
Subject 7
Trajectory C: Ipsilateral Orthogonal
Trajectory D: Contralateral Orthogonal
Subject 8
Trajectory C: Ipsilateral Orthogonal
Trajectory D: Contralateral Orthogonal

**Figure S1. Comparison of trajectories C and D.** Simulations of the most orthogonal ventral striatum trajectories are shown for Figure 3 using CT-based models in BabelBrain. Left panels show the ipsilateral orthogonal trajectory (Trajectory C); right panels show the contralateral orthogonal trajectory (Trajectory D). Each row includes a 3D head rendering from Brainsight illustrating the transducer placement (green ring), followed by 2D BabelBrain maps of the normalized acoustic intensity in the X–Z and Y–Z planes. In the 2D maps, the green line marks the ideal perpendicular (normal) angle relative to the skull surface, while the pink line indicates the actual angle at which the ultrasound beam enters the skull. A smaller deviation between the pink and green lines indicates a more orthogonal incidence. Across all subjects, Trajectory D consistently shows a beam angle closer to the surface normal than Trajectory C. The top dotted line denotes the skin surface, and the two lower dotted lines mark the skull layers. The equivalent panels are shown for Subject 1 in Figure 2.

**Figure S2.**

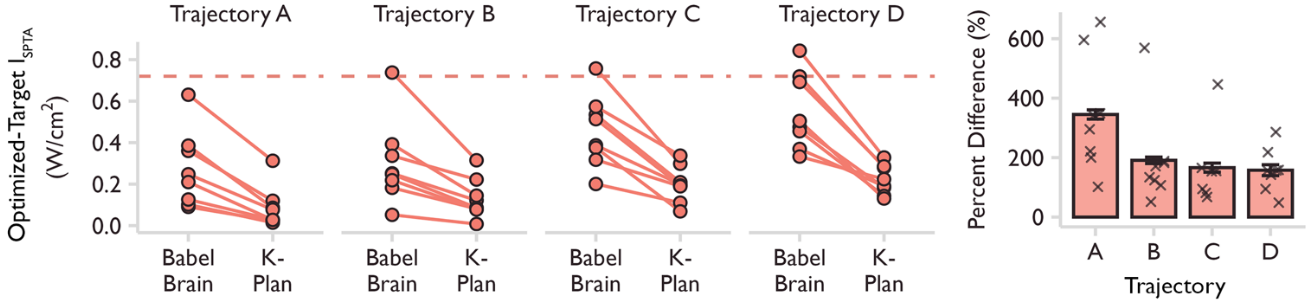


**Figure S2. $I_{SPTA}$ values from k-Plan and BabelBrain varied substantially across all trajectories for VS simulations.** Left panel: For each subject (n=8), simulation results from the four trajectories (A-D) were compared between k-Plan and BabelBrain. The dashed red line indicates the $I_{SPTA}$ safety threshold of 0.72 W/cm$^2$. All trajectories were simulated at 60 W/cm$^2$ $I_{SPPA}$ in water. Right panel: Percent differences between BabelBrain and k-Plan outputs are shown for each trajectory across all five subjects, with mean ± SEM overlaid. Percent difference was calculated as:

$$\frac{BabelBrain\ Output - k{-}Plan\ Output}{k{-}Plan\ Output}\ x\ 100.$$

**Figure S3.**

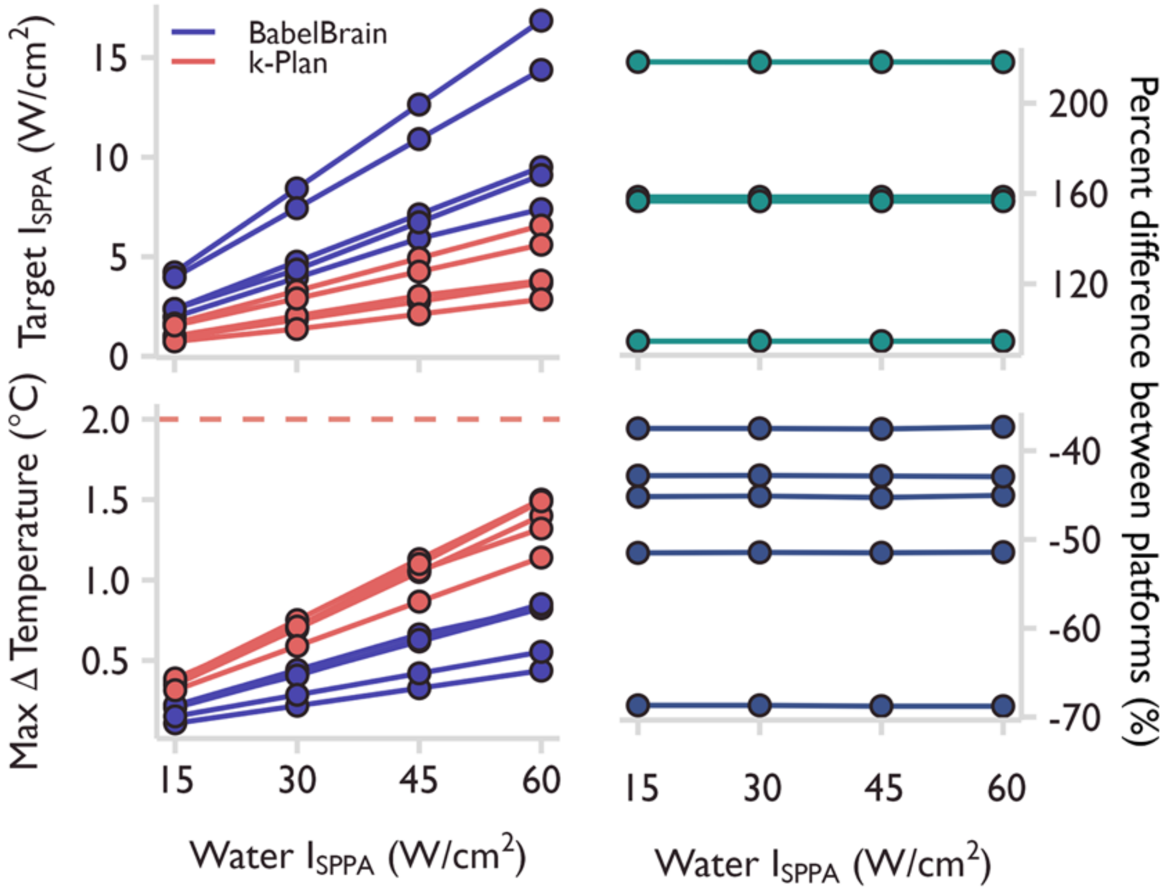


**Figure S3. Changing sonication intensity does not affect the relative difference between simulation results across simulation platforms.** Left panels: For Trajectory D across all subjects (n=5, target-derated brain $I_{SPPA}$ and the max change in skull temperature (°C) are shown for simulations at four free-field $I_{SPPA}$ in water (15, 30, 45, 60 W/cm$^2$) performed in both k-Plan and BabelBrain. Dashed red lines mark the maximum temperature rise safety threshold of 2°C. Right panels: Percent differences between BabelBrain and k-Plan outputs are shown at each intensity level for each subject. Percent difference was calculated as: $\frac{BabelBrain\ Output - k\text{-}Plan\ Output}{k\text{-}Plan\ Output} x\ 100$.

**Figure S4.**

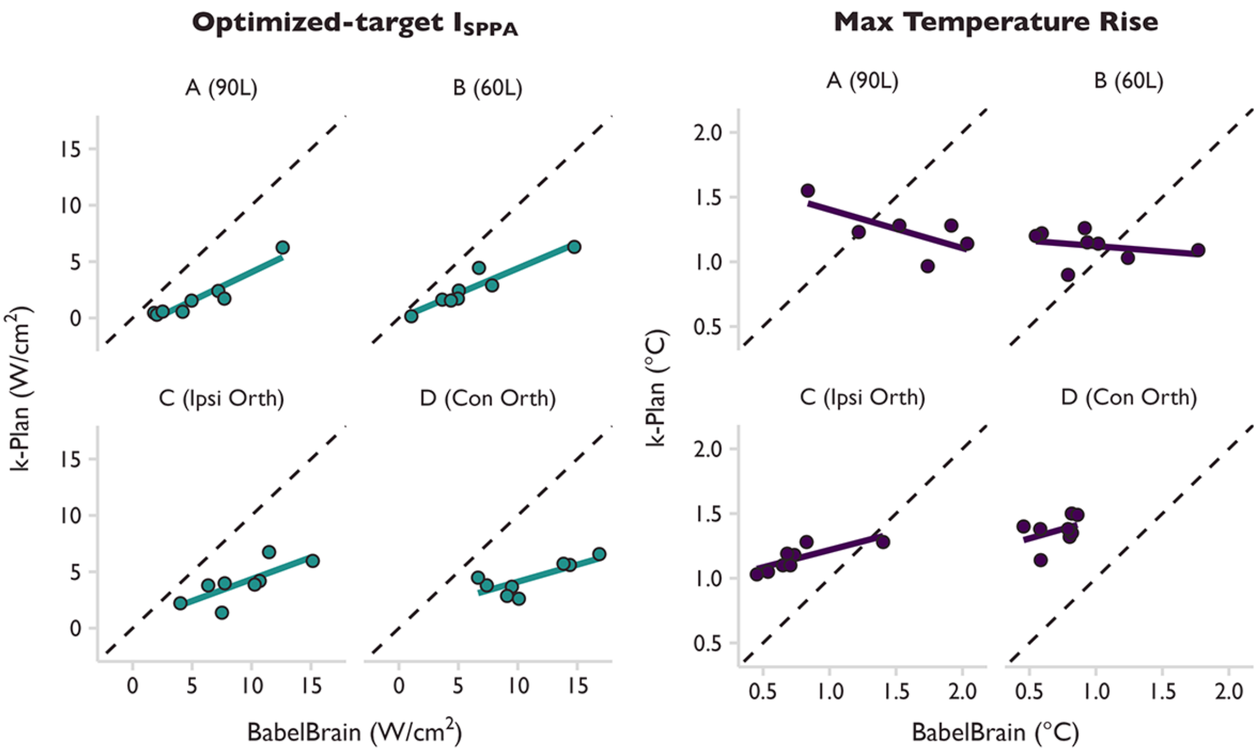


**Figure S4. Correlations between BabelBrain and k-Plan simulations.** For each subject (n=8), simulation results from the four trajectories (A-D) were correlated between k-Plan and BabelBrain for the target derated brain $I_{SPPA}$ and the max change in skull temperature (°C). All trajectories were simulated at 60 W/cm$^2$ $I_{SPPA}$ in water. Each point is a simulation from a single subject. Each panel shows the correlation for one of the four trajectories. Lines reflect linear fits between the BabelBrain and k-Plan simulations. The dashed black line indicates the identity line.

**Figure S5.**

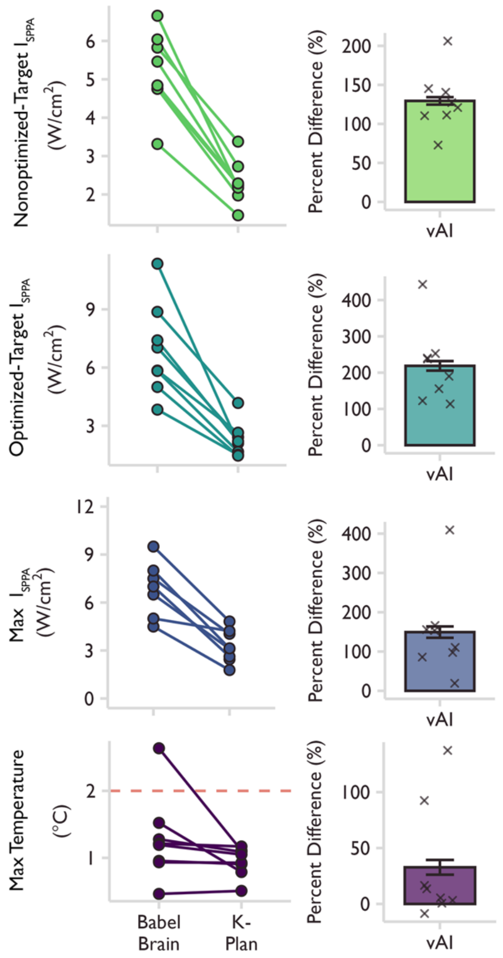


**Figure S5. Simulation outputs from k-Plan and BabelBrain varied in a second target region and using a different fundamental frequency.** Left panels: For each subject (1-8), simulation results using the CTX-250-4CH transducer to target the ventral Anterior Insula were compared between k-Plan and BabelBrain for the nonoptimized-target, optimized-target, max derated brain $I_{SPPA}$, and the max change in skull temperature (°C). The max value is the maximum $I_{SPPA}$ in the brain. The location of the max value could differ between simulation platforms, while location of the target values was the same. This trajectory was simulated at 60 W/cm² $I_{SPPA}$. Estimated focal intensities were higher in BabelBrain compared to k-Plan across the nonoptimized-target (Std. $Beta_{k\text{-}Plan>BabelBrain}$ = -1.71, 95% CI:[-2.07, -1.35], $n_{obs}$ = 16, $n_{subjects}$ = 8), the optimized-target (Std. $Beta_{k\text{-}Plan>BabelBrain}$ = -1.57, 95% CI:[-2.12, -1.02], $n_{obs}$ = 16, $n_{subjects}$ = 8), and at the maximum intensity (Std. $Beta_{k\text{-}Plan>BabelBrain}$ = -1.48, 95% CI:[-2.18, -0.77], $n_{obs}$ = 16, $n_{subjects}$ = 8). The dashed red line indicates the max change in temperature safety threshold of 2°C. As with the VS, we found that for one subject, the estimated temperature rise at the skull exceeded the safety limit in one platform, but not in the other. However, we could not detect a consistent magnitude and direction of differences for estimated temperature rise in the vAI (Std. $Beta_{k\text{-}Plan>BabelBrain}$ = -0.68, 95% CI:[-1.72, 0.36], $n_{obs}$ = 16, $n_{subjects}$ = 8). Right panels: Percent differences between BabelBrain and k-Plan outputs are shown across all eight subjects, with mean ± SEM overlaid.

Percent difference was calculated as: $\frac{BabelBrain\ Output - k\text{-}Plan\ Output}{k\text{-}Plan\ Output} x\ 100$

**Figure S6.**

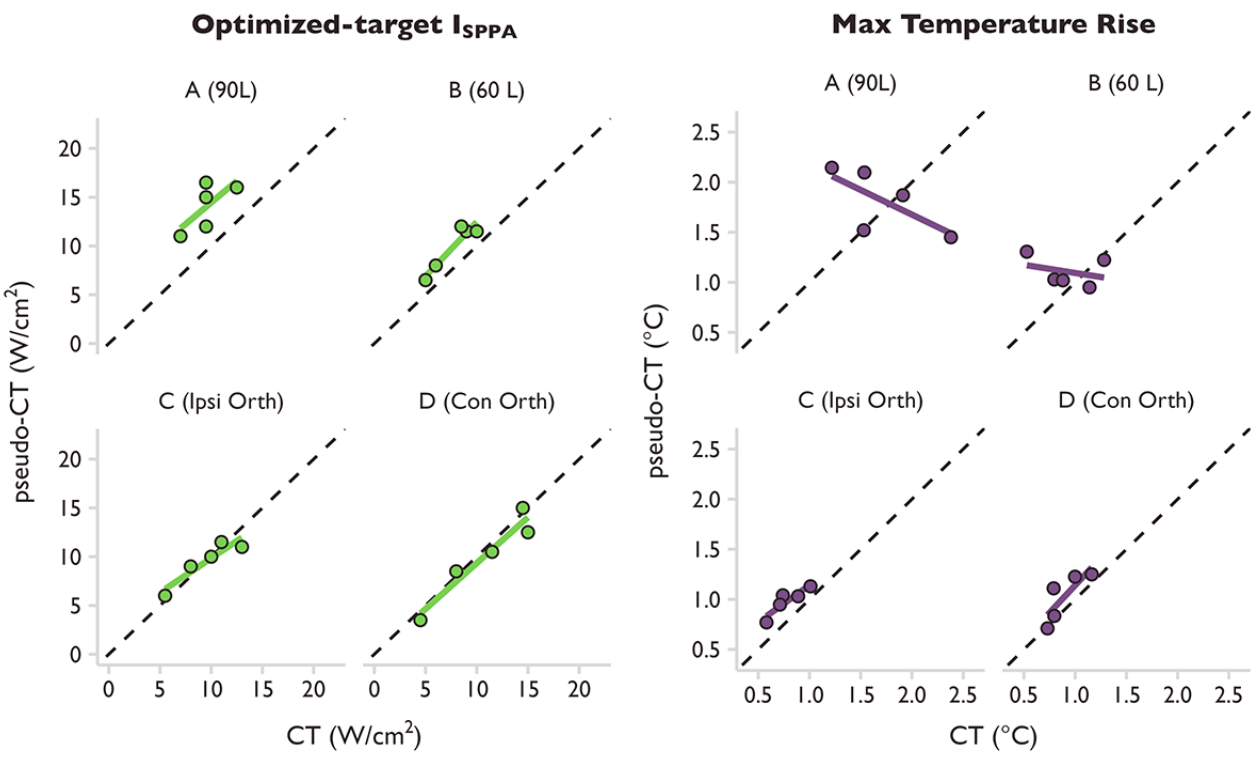


**Figure S6. Correlations between CT and pseudo-CT-based simulations.** For each subject, simulation results from the four trajectories (A-D) were correlated between the CT and pseudo-CT-based simulations for the target derated brain $I_{SPPA}$ and the max change in skull temperature (°C). All trajectories were simulated at 60 W/cm$^2$ $I_{SPPA}$ in water. Each point is a simulation from a single subject. Each panel shows the correlation for one of the four trajectories. Lines reflect linear fits between the CT and pseudo-CT-based simulations. The dashed black line indicates the identity line.

**Figure S7.**

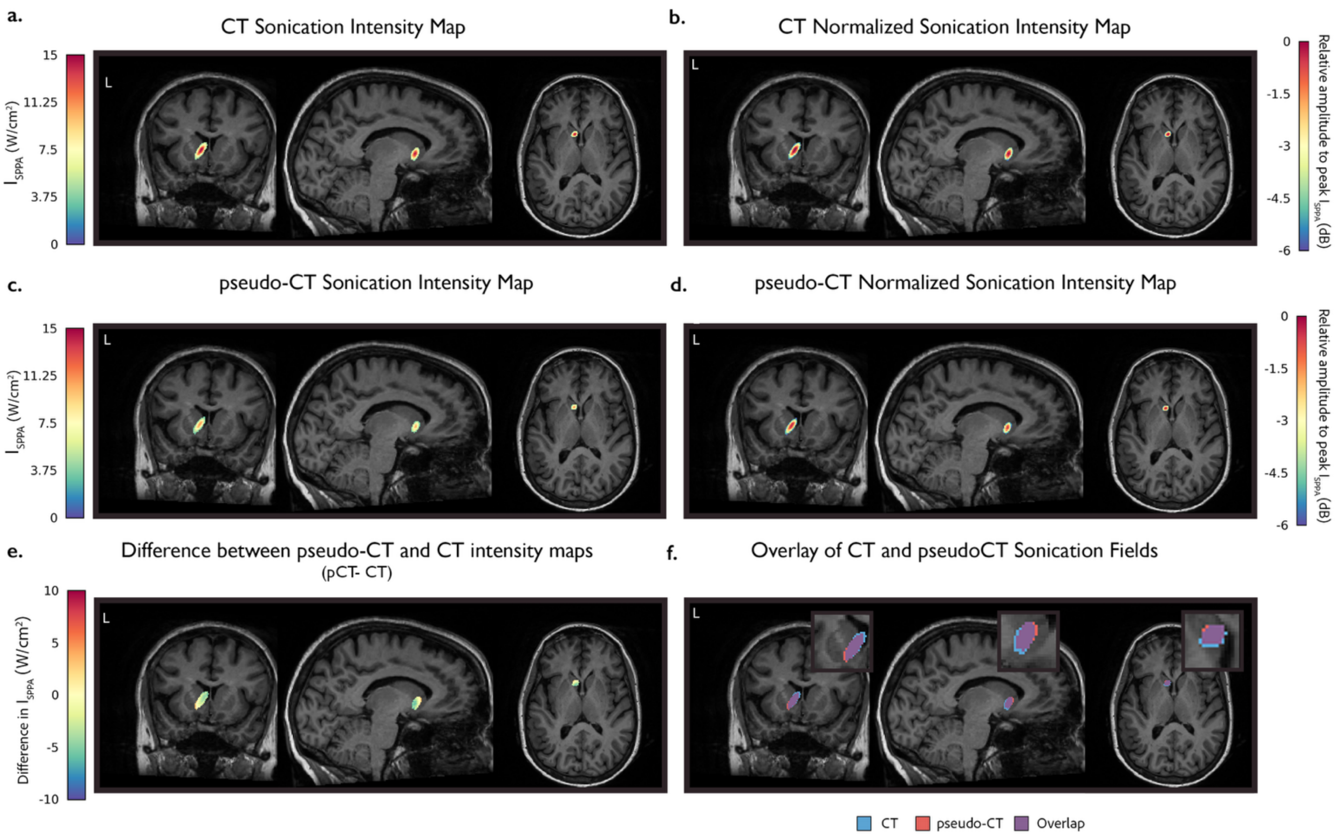


**Figure S7. Sonication beam shape and intensity comparison across CT and pseudo-CT planning images. (a–b)** CT sonication intensity maps overlaid on the subject's T1-weighted MRI. Panel **(a)** shows absolute spatial-peak pulse-average intensity ($I_{SPPA}$; W/cm²), while panel **(b)** shows intensity normalized to the peak $I_{SPPA}$ and expressed in decibels (dB). Both maps are thresholded at −6 dB (25% of peak intensity). **(c–d)** Same as panels **(a–b)**, but for the pseudo-CT sonication intensity maps. **(e)** Difference between the pseudo-CT and CT absolute intensity maps (pCT− CT). Positive (negative) values indicate regions where pseudo-CT predicts higher (lower) intensity than CT. **(f)** Spatial overlap of the −6 dB sonication regions from pseudo-CT and CT. Insets show enlarged views of the focal region near the target.

**Table S1.**

| | |
|---|---|
| **Nonoptimized-Target $I_{sppa}$** | |
| Mean ± SD | 129.5 ± 38.2% |
| Range | 72.5 – 206.0% |
| **Optimized-Target $I_{sppa}$** | |
| Mean ± SD | 218.7 ± 105.1% |
| Range | 112.4 – 441.3% |
| **$I_{sppa}$ Max** | |
| Mean ± SD | 149.3 ± 115.5% |
| Range | 18.6 – 409.2% |
| **Temp. Rise** | |
| Mean ± SD | 32.8 ± 52.8% |
| Range | -8.9 – 137.4% |

**Supplemental Table 1. Percent change between simulation platforms in intensity and temperature rise in the ventral Anterior Insula when using the CTX-250-4CH transducer.** We compared the intensity values between the simulation platforms at a free-field $I_{SPPA}$ of 60 W/cm² across three different focal points (nonoptimized-target, optimized-target, and max) and estimated max temperature rise in the skull. Mean, standard deviation (SD), and range are reported. Percent difference was calculated as:

$$\frac{BabelBrain\ Output - k\text{−}Plan\ Output}{k\text{−}Plan\ Output} x\ 100$$

**Table S2.**

| | A | B | C | D |
|---|---|---|---|---|
| *Target $I_{sppa}$* | | | | |
| Mean ± SD | 48.6 ± 20.7% | 29.5 ± 9.5% | 2.2 ± 10.9% | -7.6 ± 12.4% |
| Range | 26.3 – 73.7% | 15.0 – 41.2% | -15.4 – 12.5% | -22.2 – 6.2% |
| *Target $I_{spta}$* | | | | |
| Mean ± SD | 48.6 ± 20.7% | 29.5 ± 9.5% | 2.2 ± 10.9% | -7.6 ± 12.4% |
| Range | 26.3 – 73.7% | 15.0 – 41.2% | -15.4 – 12.5% | -22.2 – 6.3% |
| *Temp. Rise* | | | | |
| Mean ± SD | 14.1 ± 43.7% | 34.2 ± 65.6% | 26.9 ± 12.3% | 14.6 ± 17.2% |
| Range | -39.1 – 76.0% | -16.7 – 147.3% | 11.9 – 40.6% | -2.7 – 40.5% |

**Table S2. Percent change in simulation outputs when using the pseudo-CT versus CT as the planning image across four trajectories.** Simulations were computed at a free-field $I_{SPPA}$ of 60 W/cm². Mean, standard deviation (SD), and range are reported. Percent difference was calculated as:

$$\frac{pCT\ Output - CT\ Output}{CT\ Output} x\ 100$$
